# Increased frequency of repeat expansion mutations across different populations

**DOI:** 10.1101/2023.07.03.23292162

**Authors:** Kristina Ibañez, Bharati Jadhav, Matteo Zanovello, Delia Gagliardi, Christopher Clarkson, Stefano Facchini, Paras Garg, Alejandro Martin-Trujillo, Scott J Gies, Valentina Galassi Deforie, Anupriya Dalmia, Davina J. Hensman Moss, Jana Vandrovcova, Clarissa Rocca, Loukas Moutsianas, Chiara Marini-Bettolo, Helen Walker, Chris Turner, Maryam Shoai, Jeffrey D Long, EUROSCA network, Pietro Fratta, Douglas R Langbehn, Sarah J Tabrizi, Mark J Caulfield, Andrea Cortese, Valentina Escott-Price, John Hardy, Henry Houlden, Andrew J Sharp, Arianna Tucci

## Abstract

Repeat expansion disorders (REDs) are a devastating group of predominantly neurological diseases. Together they are common, affecting 1 in 3,000 people worldwide with population-specific differences. However, prevalence estimates of REDs are hampered by heterogeneous clinical presentation, variable geographic distributions, and technological limitations leading to under-ascertainment. Here, leveraging whole genome sequencing data from 82,176 individuals from different populations, we found an overall disease allele frequency of REDs of 1 in 283 individuals. Modelling disease prevalence using genetic data, age at onset and survival, we show that the expected number of people with REDs would be two to three times higher than currently reported figures, indicating under-diagnosis and/or incomplete penetrance. While some REDs are population-specific, e.g. Huntington disease-like 2 in Africans, most REDs are represented in all broad genetic ancestries (i.e. Europeans, Africans, Americans, East Asians, and South Asians), challenging the notion that some REDs are found only in specific populations. These results have worldwide implications for local and global health communities in the diagnosis and counselling of REDs.

## INTRODUCTION

Repeat expansion disorders (REDs) are a heterogeneous group of conditions which mainly affect the nervous system, and include Fragile X syndrome, the commonest inherited form of amyotrophic lateral sclerosis and frontotemporal dementia (*C9orf72*-ALS/FTD)^1^, and inherited ataxias (Friedreich ataxia (FA), and *RFC1*-Cerebellar Ataxia, Neuropathy, Vestibular Areflexia syndrome, CANVAS^2^). REDs are caused by the same underlying mechanism: the expansion of short repetitive DNA sequences (1-6 bp) within their respective genes. The mutational process is gradual: normal alleles are usually passed stably from parent to child with rare changes in repeat size; intermediate-size are more likely to expand into the disease range, giving rise to pathogenic repeat lengths in the next generation. Repeat lengths are classified in ascending order as normal, intermediate, premutation, reduced penetrance, or full mutations, though this classification is not universal, and not all RED loci have well-defined ranges for intermediate or reduced penetrance range.

REDs are clinically heterogeneous: for example, *C9orf72* expansions can present as either FTD or ALS even within the same family; and one in three patients carrying the repeat expansion in *C9orf72* shows an atypical presentation at onset such as Alzheimer’s and Huntington’s disease (HD) among others^3,4^. For many REDs, the variability in repeat lengths underlines the substantial clinical heterogeneity^5^, longer repeats cause more severe disease, and earlier symptom onset^6^.

Previous studies have estimated that REDs affect 1 in 3,000 people^7^. Despite their broad distribution in human populations, few global epidemiological studies have been performed. In these studies, prevalence estimates are either population-based, in which affected individuals are identified based on clinical presentation, or genetically tested based on the presence of a relative with a RED. Given that one of the most striking features of REDs is that they can present with markedly diverse phenotypes, REDs can remain unrecognised, leading to underestimation of the disease prevalence^8^.

While many of the epidemiological studies to date have been conducted in cohorts of European origin, studies in other ancestries have highlighted population differences at specific RED loci^9–11^. Among the most common REDs, myotonic dystrophy type 1 (DM1) affects 1 in 8,000 people worldwide^12^, ranging from 1 in 10,000 in Iceland to 1 in 100,000 in Japan^13^. Similarly, HD prevalence ranges from 0.1 in 100,000 in Asian and African countries^14,15^ to 10 in 100,000 in Europeans^16^. In Europeans, it is estimated that the prevalence of *C9orf72*-FTD is 0.04-134 in 100,000, and *C9orf72*-ALS is 0.5-1.2 in 100,000^4^. The spinocerebellar ataxias (SCAs) are a group of rare neurodegenerative disorders mainly affecting the cerebellum. They are individually rare worldwide, with largely variable frequencies among populations^17^, mainly due to founder effects. Overall, the worldwide prevalence of SCAs is 2.7-47 cases per 100,000^10^ with SCA3 being the most common form worldwide, followed by SCA2, SCA6, and SCA1^18^.

With the advent of disease-modifying therapies for REDs, it is becoming necessary to determine comprehensively the number of patients and type of RED expected in different populations, so that targeted approaches can be developed accordingly. Large-scale genetic analyses of REDs have been limited by repeat expansion profiling techniques, which historically have relied on polymerase chain reaction-based (PCR) assays or Southern blots, which by nature are targeted assays and can be difficult to scale. So far, the largest population study of the genetic frequency REDs involved the PCR-based analysis of 14,196 individuals of European ancestry^19^.

In the last few years, bioinformatic tools have been developed to profile DNA repeats from short-read whole exome^20^ and whole genome sequencing data (WGS)^21^. We have recently shown that disease-causing repeat expansions can be detected from WGS with high sensitivity and specificity, making large-scale WGS datasets an invaluable resource for the analysis of the frequency and distribution of REDs^7^. Our group has previously applied this pipeline to a large WGS cohort to assess the distribution of repeat expansions in the *AR* gene, which cause Spinal and Bulbar Muscular Atrophy (SBMA), and found an unexpectedly high frequency of pathogenic alleles, suggesting under-diagnosis or incomplete penetrance of this RED^22^. However, a comprehensive study of REDs in the general population and across different ancestries using WGS has never been performed.

In the present study, we used large-scale genomic databases to address two main questions: i) what is the frequency of RED mutations in the general population? ii) how does the frequency and distribution of REDs vary across populations?

## RESULTS

### Cohort description

We analysed RED loci from two large-scale medical genomics cohorts with high-coverage WGS and rich phenotypic data: the 100,000 Genomes Project (100K GP) and Trans-Omics for Precision Medicine (TOPMed). The 100K GP is a programme to deliver genome sequencing of people with rare diseases and cancer within the National Health Service (NHS) in the United Kingdom^23,24^. TOPMed is a clinical and genomic programme focused on elucidating the genetic architecture and risk factors of heart, lung, blood, and sleep disorders from the National Institutes of Health (NIH)^25^. First, we selected WGS data generated using PCR-free protocols and sequenced with paired-end 150 bp reads (**Table S1, Methods**). To avoid overestimating the frequency of REDs, we excluded individuals with neurological diseases, as their recruitment was driven by the fact that they had a neurological disease potentially caused by a repeat expansion. We then performed relatedness and principal component analyses to identify a set of genetically unrelated individuals and predict broad genetic ancestries based on 1,000 Genomes Project super-populations^26^ (1K GP3). The resulting dataset comprised a cross-sectional cohort of 82,176 genomes from unrelated individuals (median age 61, Q1-Q3: 49-70, 58.5% females, 41.5% males; **Table S2, Extended data Fig.1**), genetically predicted to be of European (n=59,568), African (n=12,786), American (n=5,674), South Asian (n=2,882), and East Asian (n=1,266) descent (**Extended data Fig. 2, Methods**).

### RED mutation frequency

To estimate the number of individuals carrying premutation or full mutation alleles (**Table 1**), we selected repeats in RED genes^27^ for which WGS can accurately discriminate between normal and pathogenic alleles^7^, based on either or both, of the following conditions: the threshold between premutation and full mutation is shorter than the sequencing read-length (and therefore WGS can accurately distinguish between premutation and full mutation), or WGS was validated against the current gold-standard PCR test (**Extended data Fig. 3**, **Table S3**). For the latter, PCR tests were obtained from a cohort of individuals recruited to the 100K GP who had RED testing as part of their standard diagnostic pathway (**Table S4, Methods**). Within this dataset, we show that: i) WGS accurately classifies alleles in the normal, premutation, and full mutation range in all loci assessed except *FMR1* (that causes Fragile X syndrome) (**Extended data Fig. 3A, Table S4)**; ii) the accuracy of repeat sizing by WGS is not affected by genetic ancestry by comparing genotypes generated by WGS to PCR from different populations (**Extended data Fig. 3B**) but it might underestimate the size of large expansions in *FMR1*, *DMPK*, *FXN*, and *C9orf72,* as previously described^7^.

Table 1

**Table 1**
| Disease | Locus | Premutation/reduced penetrance | Full Mutation repeat threshold |
| --- | --- | --- | --- |
| Spinal and bulbar muscular atrophy | *AR* | 35-37 | 38 |
| Dentatorubral-pallidoluysian atrophy | *ATN1* | 35-47 | 48 |
| Spinocerebellar ataxia 1 | *ATXN1* | 39-43 | 44 |
| Spinocerebellar ataxia 2 | *ATXN2* | 33-34 | 35 |
| Spinocerebellar ataxia 3 | *ATXN3* | 45-59 | 60 |
| Spinocerebellar ataxia 7 | *ATXN7* | 34-36 | 37 |
| C9orf72-related Frontotemporal dementia and/or amyotrophic lateral sclerosis 1 | *C9orf72* | - | 31 |
| Spinocerebellar ataxia 6 | *CACNA1A* | 19 | 20 |
| Myotonic dystrophy 1 | *DMPK* | 35-49 | 50 |
| Friedreich ataxia | *FXN* | 45-65 | 66 |
| Huntington disease | *HTT* | 36-39 | 40 |
| Huntington disease-like 2 | *JPH3* | 29-39 | 40 |
| Oculopharyngodistal myopathy 3; Neuronal intranuclear inclusion disease (NIID) | *NOTCH2NLC* | 41-55 | 56 |
| Spinocerebellar ataxia 12 | *PPP2R2B* | 32-65 | 66 |
| Cerebellar ataxia, neuropathy, and vestibular areflexia syndrome | *RFC1* | - | 1042 (median number of repeats)<sup>56</sup><br>IQR=844-1306<br>range=249-3885 |
| Spinocerebellar ataxia 17 | *TBP* | 41-48 | 49 |

Furthermore, as we previously developed and validated a dedicated WGS analytical workflow for the repeat expansion in *RFC1* that causes CANVAS^28^, this repeat was included in our analysis.

Overall, 16 RED loci pass our criteria for accurately estimating premutation and full mutation carrier frequencies, representing a broad spectrum of REDs and different modes of inheritance: 1) autosomal dominant: HD, Huntington disease-like 2 (HDL2), DM1, *C9orf72*-ALS/FTD, the spinocerebellar ataxias (SCA1, SCA2, SCA3, SCA6, SCA7, SCA12, SCA17), dentatorubral-pallidoluysian atrophy (DRPLA), and *NOTCH2NLC*, which causes a spectrum of neurological disorders, especially neuronal intranuclear inclusion disease and oculopharyngodistal myopathy; 2) autosomal recessive: Friedreich ataxia and CANVAS, 3) X-linked SBMA (**Table 1**).

Our analysis workflow (**Fig. 1**) included profiling each RED locus, followed by quality control of all alleles (employing Expansion Hunter classifier, https://github.com/bharatij/ExpansionHunter_Classifier, and visual inspection of pileup plots as previously described^29^) predicted to be larger than the premutation threshold (**Table S5, Methods**). We also retrospectively analysed factors potentially leading to overestimating disease allele frequency, such as checking that there was no selection bias for patients with DM1 which can cause cardiac abnormalities (**Table S6**).

**Figure 1.**
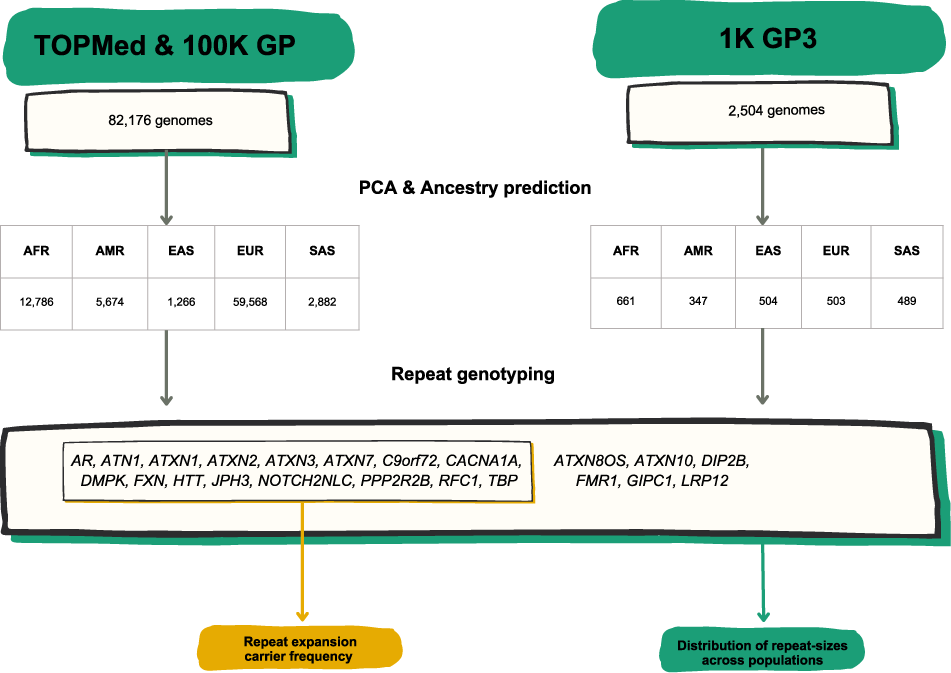
**A)** List of RED loci included in the study including repeat-size thresholds for reduced penetrance and full mutations. **B)** Technical flowchart. Whole genome sequences (WGS) from the 100K GP and TOPMed datasets were first selected by excluding those associated with neurological diseases. WGS data from the 1K GP3 were also selected by having the same technical specifications (see **Methods**). After inferring ancestry prediction, repeat sizes for all 22 REDs were computed by using EH v3.2.2. On one hand, for 16 REDs overall carrier frequency, disease modelling, and correlation distribution of long normal alleles were computed in the 100K GP and TOPMed projects. On the other hand, the distribution of repeat sizes across different populations was analysed in the 100K GP and TOPMed combined, and in the 1K GP3 cohorts.

In total, for autosomal dominant and X-linked REDs, there were 290 individuals carrying one fully expanded repeat and 1,279 individuals carrying one repeat in the premutation range, meaning that the frequency of individuals carrying full-expansion and premutation alleles among this large cohort is 1 in 283 and 1 in 64, respectively (**Table S7**).

The most common expansions (in the full-mutation range, Table 1) were those in *C9orf72* (*C9orf72*-ALS/FTD) and *DMPK* (DM1) with a frequency of 1 in 839 and 1 in 1,786 respectively, followed by expansions in *AR* (SBMA: 1 in 2,561 males) and *HTT* (HD: 1 in 4,109). Surprisingly, many individuals were found to carry expansions in the SCA genes: 1 in 5,136 in *ATXN2* (SCA2), 1 in 5,136 in *CACNA1A* (SCA6), and 1 in 6,321 in *ATXN1* (SCA1). By contrast, expansions in *ATXN7* (SCA7) and *TBP* (SCA17) were present in only two individuals at each locus (1 in 41,077, and expansions in *JPH3* (Huntington disease-like 2) and *ATN1* (DRPLA) were very rare, with only a single individual at each locus identified with a repeat allele in the pathogenic full-mutation range. No pathogenic full-mutation expansions were identified in *ATXN3* (SCA3)*, PPP2R2B* (SCA12), and *NOTCH2NLC* (**Fig. 2, Table S7**).

**Figure 2.**
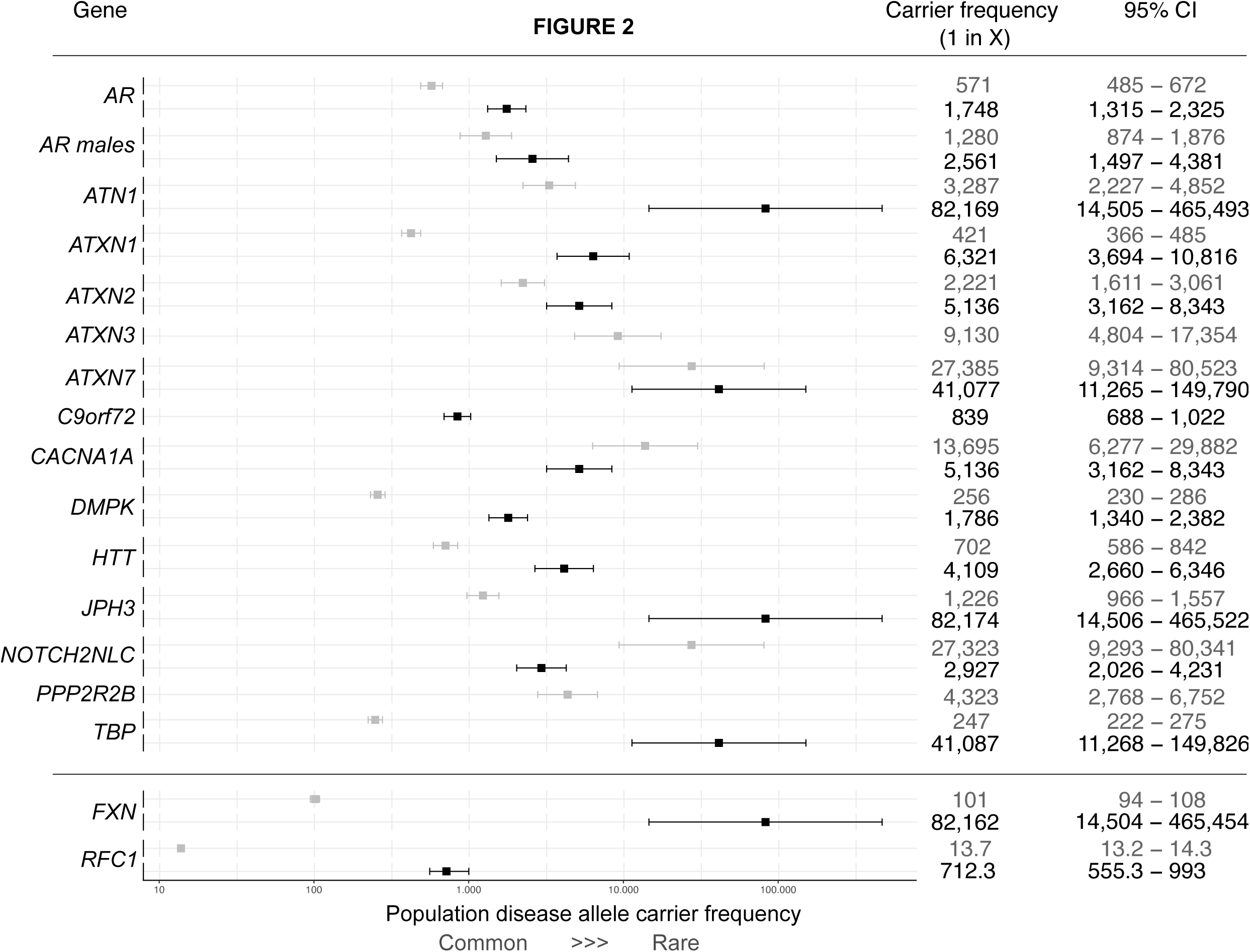
Forest plot with combined overall disease allele carrier frequency in the combined 100K GP and TOPMed datasets N = 82,176 (N individuals may vary slightly between loci due to data quality and filtering, See Table S7). The squares show the estimated disease allele carrier frequency, and the bars show the 95% CI values. Details of the statistical models are described in the Methods section. Grey and black boxes show premutation/reduced penetrance and full mutation allele carrier frequencies for each dominant locus, respectively. Grey and black boxes show mono-and bi-allelic carrier frequencies for recessive loci (*RFC1* and *FXN*), respectively.

For autosomal recessive REDs, we found a carrier frequency (i.e. people that carry one expanded allele) of 1 in 101 for *FXN* (FA) and 1 in 14 for *RFC1*; and a frequency of biallelic expansions of 1 in 82,176 for *FXN* (FA) and 1 in 712 for *RFC1* CANVAS **(Fig. 2, Table S8)**. Demographic data available on all individuals carrying a pathogenic full-mutation repeat are listed in **Table S9.** The distribution of repeat sizes overall in this cohort is represented in **Extended data Fig. 4**.

### Modelling the expected number of people affected by REDs

REDs have variable age at onset, disease duration, and penetrance^1^. Therefore, the mutation frequency cannot be directly translated into disease frequency (i.e. prevalence). To estimate the expected number of people affected by REDs, we took the mutation frequency of the most common REDs (*C9orf72*-ALS/FTD, DM1, HD, SCA1, SCA2, SCA6), and modelled the distribution by age of those expected to be affected by REDs in the UK population. For this analysis, we used the data from the Office of National Statistics^30^, and age of onset, penetrance, and impact on survival of each RED based on either cohort studies or disease-specific registries (**Methods**).

We estimated on average a two-to three-fold increase in the predicted number of people with REDs, compared to currently reported figures based on clinical observation, depending on the RED (**Fig. 3**). Since *C9orf72* expansions cause both ALS and FTD, we modelled both diseases separately, providing for *C9orf72*-ALS an expected number of people affected over two times higher than previous estimates (**Table S10**: 1.8 per 100,000 versus 0.5-1.2 in 100,000^4,31^) and for *C9orf72*-FTD 6.5 per 100,000 (**Table S11**) within the wide reported range^32,33^). For DM1, we estimated that 15.9 per 100,000 people would be affected by the condition (**Table S12**), 1.3 times higher than the estimated prevalence from clinical data (12.25 in 100,000^34^). For HD, the majority of individuals with a pathogenic expansion in our cohort carries alleles with 40 repeats (12 out of 20 people, **Table S9**). Given the well-established relationship between *HTT* repeat length and age at onset, we modelled the expected number of people with HD based on the observed frequency of the expansion, taking into account age at onset distribution and penetrance data for repeat length equal to 40 units^6^. We found that 2.3 per 100,000 people are estimated to have HD caused by 40 CAG repeats (**Table S13**): over 3 times higher than the reported number of affected patients with 40 CAG repeats (0.72 per 100,000; **Methods** and personal communication, DRL and DHM). For SCA2 and SCA1 our model indicates over three-fold increase in the number of people expected with the disease compared to the reported prevalence (3 and 3.7 per 100,000 respectively based on our estimate in **Table S14**, **Table S15** versus the currently reported prevalence of 1 per 100,000)^35,36^. Strikingly, we found that the expected number of people with SCA6 would be nine times higher than the reported prevalence: 9 in 100,000 versus 1 in 100,000 individuals (**Table S16**, **Methods**). Overall, these data indicate that REDs are either underdiagnosed or that not all individuals who carry a repeat larger than the established full mutation cut-off develop the condition (i.e. incomplete penetrance).

**Figure 3.**
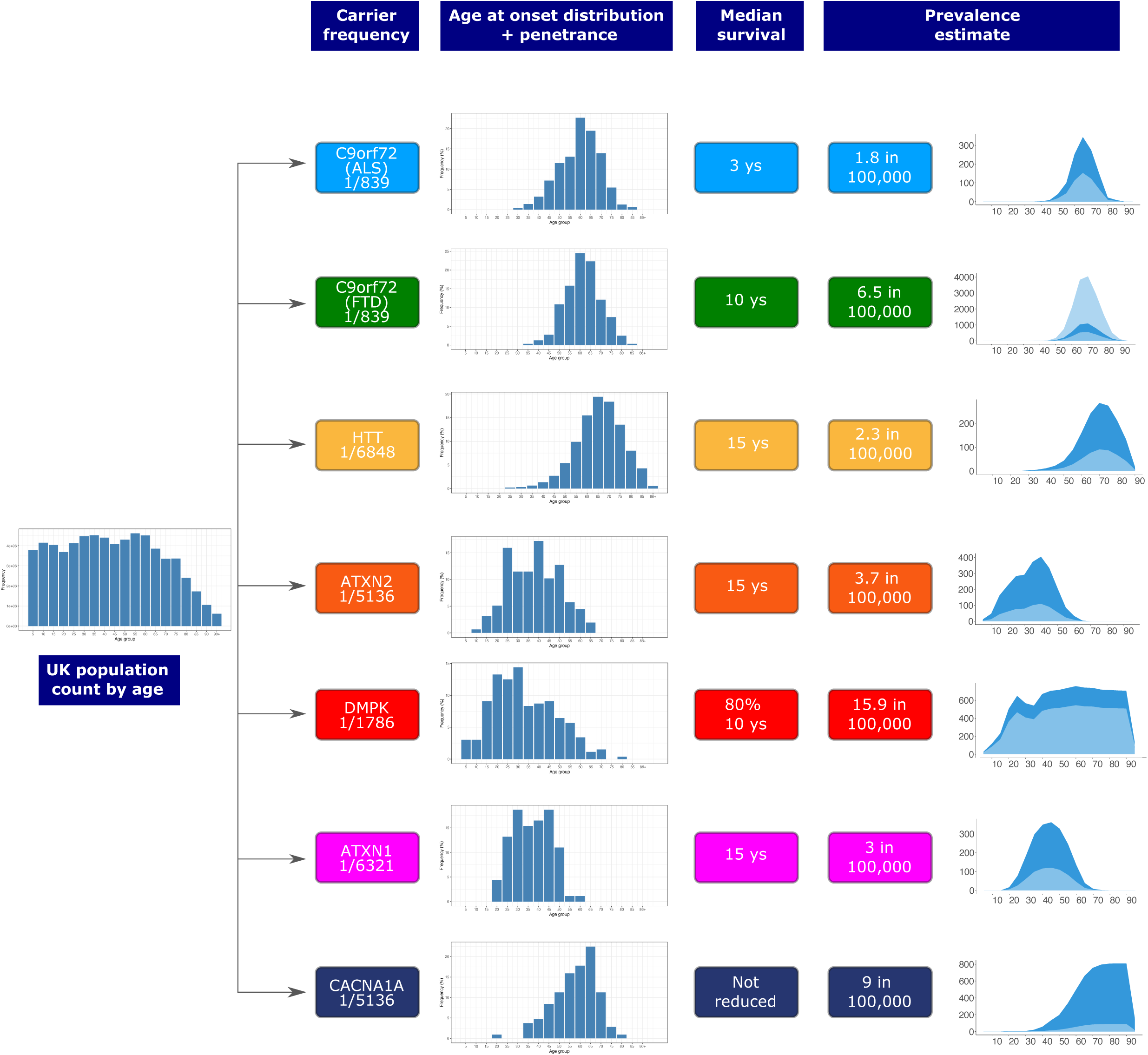
Flowchart showing the modelling of disease prevalence by age for *C9orf72*-ALS, *C9orf72*-FTD, HD in 40 CAG repeat carriers, SCA2, DM1, SCA1, and SCA6. UK population count by age is multiplied by the combined disease allele frequency of each genetic defect and the age of onset distribution of each corresponding disease, and corrected for median survival. Penetrance is also taken into account for *C9orf72*-ALS and *C9orf72*-FTD. Estimated number of people affected by REDs (dark blue area) compared to the reported prevalence from the literature (light blue area). Age bins are 5 years each. For *C9orf72*-FTD, given the wide range of the reported disease prevalence,^32,33^ both lower and upper limits are plotted in light blue.

### RED mutation frequency in different populations

The prevalence of individual REDs varies considerably based on geographic location. Hence, we set out to analyse whether these differences are reflected in the broad genetic ancestries of our cohort. First, we visualised the individuals carrying an expansion in a RED gene on the principal component analysis plot (**Extended data Fig. 5**). We then computed the proportion of pathogenic allele carriers (premutation and full mutation allele carriers) in each population (**Fig. 4A, Table S17**). In agreement with current known epidemiological studies, we observed that pathogenic alleles in *FXN* (FA), *C9orf72*, and *DMPK* (DM1) are more common in European; those in *ATN1* (DRPLA)*, TBP* (SCA17), and in *NOTCH2NLC* are more common in East Asians, and those in *JPH3* (HDL2) are more common in Africans. Conversely, pathogenic alleles in *ATXN2* are more equally distributed across different populations, and those in *RFC1* are less prevalent in Africans. Moreover, pathogenic expansions within *C9orf72* and *HTT* were identified in Africans and South Asians, which to date have only been reported in smaller clinical studies^11,37–40^. Given that the initial ancestry assignments for our cohort were based on genome-wide data, we performed local ancestry analysis to check for admixture in these individuals, confirming that the expanded repeat alleles segregated on haplotypes of African and South Asian ancestry (**Methods**).

**Figure 4.**
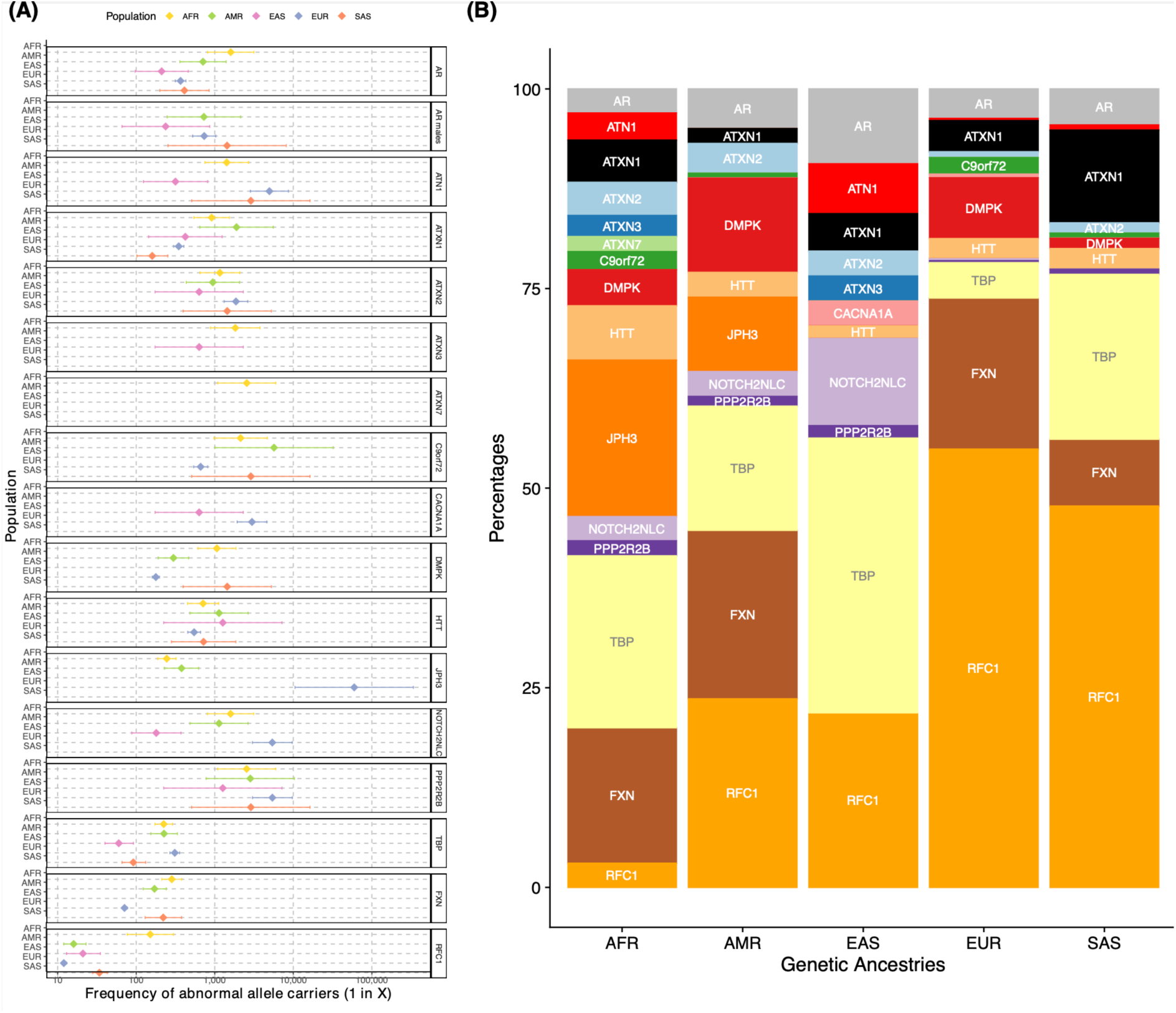
Pathogenic RED frequencies in different populations (African = 12,786; American = 5,674; East Asian = 1,266; European = 59,568; South Asian = 2,882. **A)** Forest plot of pathogenic allele carrier frequency divided by population. Pathogenic alleles are defined as those larger than the premutation cut-off (Table1). Data are presented as squares showing the estimated pathogenic allele carrier frequency, and bars showing the 95% CI values. **B)** Bar chart showing the proportion of pathogenic allele carrier frequency repeats by ancestry. Both plots have been generated by combining data from 100K GP and TOPMed from a total of N = 82,176 unrelated genomes. N individuals may vary slightly between loci due to data quality and filtering, see **Table S17** and **Table S18**).

**Figure 5.**
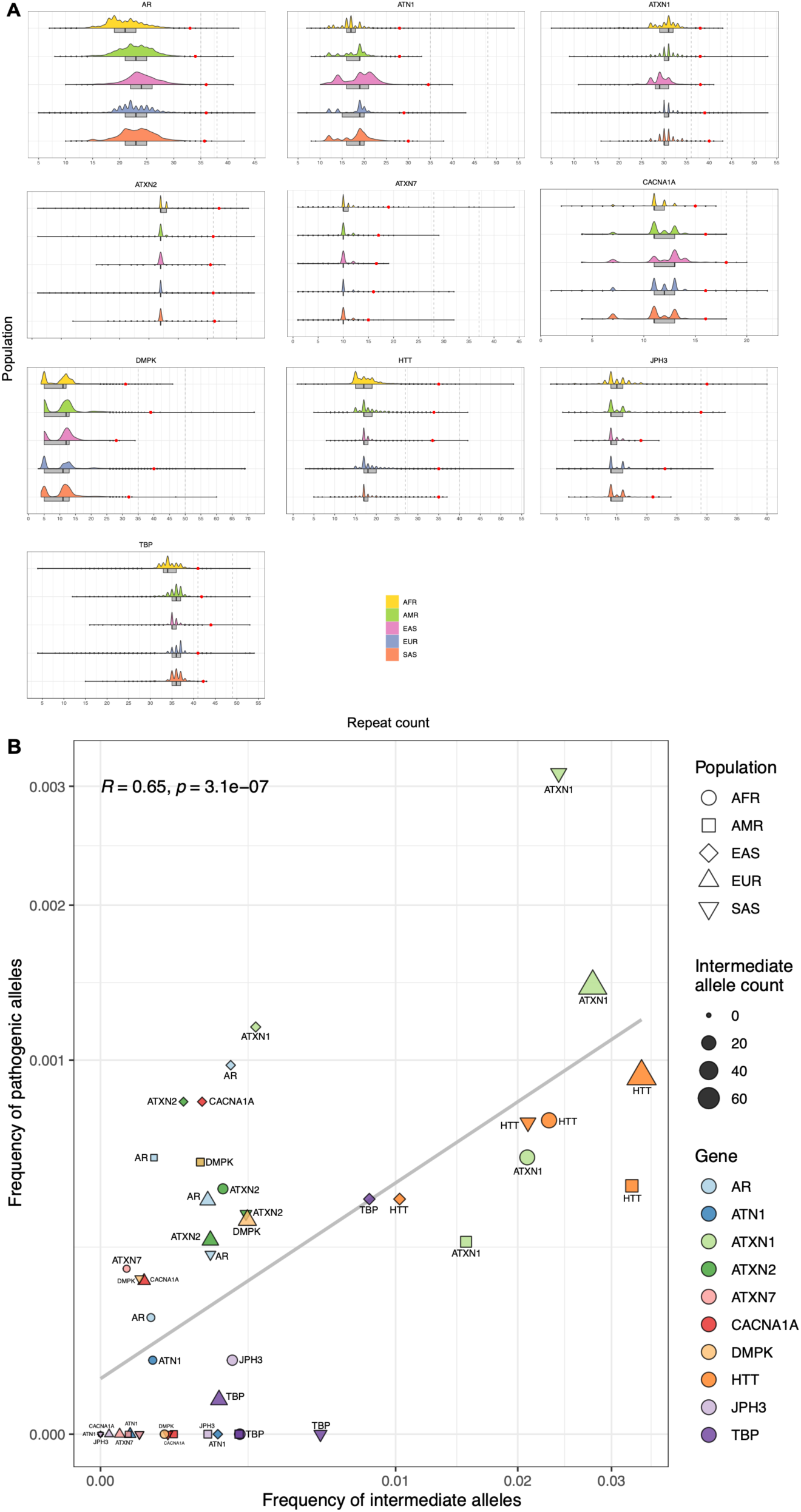
Distribution of repeat lengths in different populations. **A)** Half-violin plots showing the distribution of alleles in different populations (African = 12,786; American = 5,674; East Asian = 1,266; European = 59,568; South Asian = 2,882) for 10 loci (**Methods**) from the combined 100K GP and TOPMed cohorts. Box plots highlight the interquartile range and median, and black dots show values outside 1.5 times the interquartile range. Red dots mark the 99.9^th^ percentile for each population and locus. Vertical bars indicate the intermediate and pathogenic allele thresholds (**Table S20**). **B)** Scatter plot shows the frequency of intermediate allele carriers (x-axis) against the frequency of pathogenic allele carriers (y-axis). Data points are divided by population (n=5) and gene (n=10), and size represents the total number of intermediate alleles. Correlations were computed using the Spearman method, and two-tailed p-values.

We then analysed the relative frequency of pathogenic allele carriers within each population (**Fig. 4B, Table S18**), highlighting differences in the proportion of REDs within and between populations. Pathogenic allele carriers were observed in most REDs across all populations (e.g. those in *AR*, *ATXN1*, *ATXN2*, *HTT* and *TBP*), though in variable proportions and with some notable exceptions. Pathogenic alleles in *RFC1* are by far the most widely represented (followed by those in *AR* and *TBP*). The African population is the most diverse, with pathogenic expansions present across all RED loci, except *CACNA1A*. The East Asian population is the one with more striking differences in the relative frequency of REDs, and notably the absence of pathogenic alleles in *FXN* and *DMPK* and the large proportion of *TBP* (signal driven mainly reduced penetrance alleles; **Table S7** shows that the carrier frequency of reduced penetrance alleles).

### Distribution of repeat lengths in different populations

REDs are thought to arise from large normal polymorphic repeats (large normal, or “intermediate” range repeats), as they have an increased propensity to further expand upon transmission from parent to progeny, moving into the pathogenic range. The uneven RED prevalence across major populations has been associated with the variable frequency of intermediate alleles^41,42^.

Therefore, we analysed intermediate allele frequencies for those genes where WGS can accurately size intermediate alleles (**Methods**) across populations and confirmed that: i) the overall distribution of repeat lengths varies across populations **(Fig. 5A, Extended data Fig. 6, Table S19)**; for example, the median repeat size of *PPP2R2B* is higher in East and South Asians compared to Europeans (13 versus 10 repeats); ii) overall, the frequency of intermediate alleles varies in each population, and correlates with the frequency of pathogenic alleles; (R = 0.65; p = 3.1×10^-7^, Spearman correlation) (**Fig. 5B**, **Extended data Fig. 7, Table S20**). These data suggest that different distributions of repeat lengths underlie differences in the epidemiology of REDs.

In fact, for HD^43,44^, specific structures have been proposed as *cis*-acting modifiers of HD^39^. In a typical *HTT* allele, the pure CAG tract (Q1) is followed by an interrupting CAACAG sequence (Q2). These are followed by a polyproline region encoded by a CCGCCA sequence (P1), then by stretches of CCG repeats (P2), and, lastly, by CCT repeats (P3) (**Fig. 6A**). Variations in this sequence have been described, including duplication or loss of Q2 or loss of P1, and variation in the number of the downstream P2 and P3 repeats. To analyse population differences in the repeat structures of *HTT*, we developed an analytical workflow to determine accurately phased Q1, Q2, and P1 elements of the *HTT* structure from WGS (**Methods)**. We confirm that the typical structure Q1_n_-Q2_1_-P1_1_ (also known as ‘canonical’) is the most common across populations, and that other structures are present in different proportions in different populations: P1 loss alleles are more prevalent in Africans and East Asians; while overall non-canonical alleles are more frequent in African, p < 0.0001 two-tailed Chi-square test) (**Fig. 6B**). Moreover, variable Q1 repeat lengths are associated with different structures (linear model, p-value <2.2×10^-16^): shorter Q1 lengths are found on chromosomes with Q2 duplication, and larger Q1 lengths are found in those with Q2 loss and Q2-P1 loss (**Fig 6C**).

**Figure 6.**
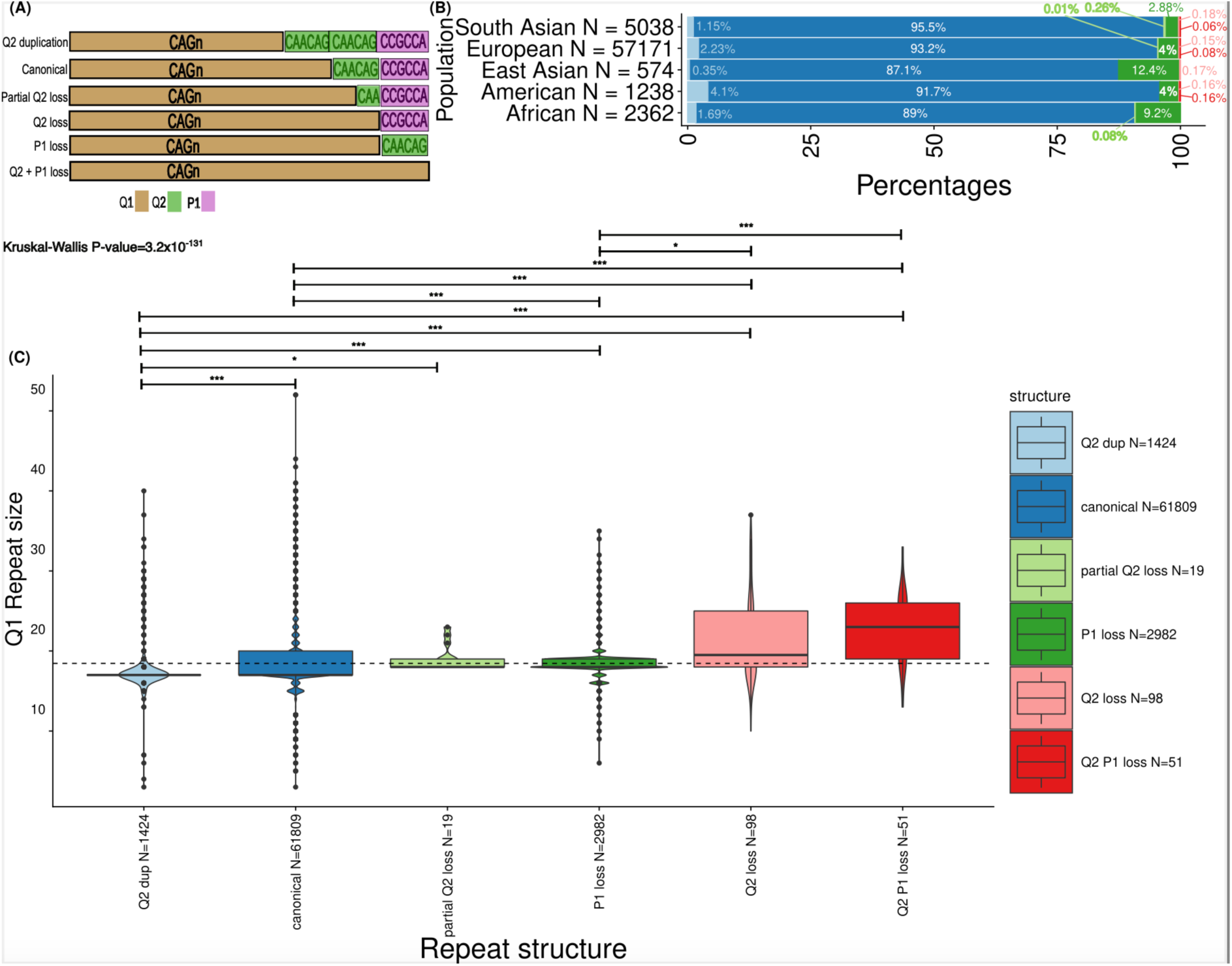
*HTT* repeat structures show varied prevalence across genetic ancestries and are associated with CAG repeat size. **A)** Allele structures observed within exon 1 of *HTT*. The CAG repeat is denoted as “Q1” and marked in gold. The CAACAG unit is referred to as “Q2” and is marked in green. The first proline-encoding “CCGCCA” repeat element is referred to as “P1” and is marked in purple. **B)** The prevalence of the allele structures is plotted across the studied genetic ancestries in bar plots on the x-axis. The ancestries are defined on the y-axis. The number of alleles in each of the genetic ancestries is denoted as “N=…” at each of the y-axis ticks. **C)** Boxplots display the distribution of CAG repeat sizes across different repeat structures. Box plots highlight the median (horizontal lines in the centre of each boxplot), interquartile range (bounds) and black dots show values outside 1.5 times the interquartile range. The repeat structures are separated on the x-axis and the repeat size is shown on the y-axis. The number of alleles with different repeat structures is denoted as “N=…” on the x-axis. A linear model was used to compare the repeat size distribution of the canonical alleles versus that of all atypical structures. Kruskal-Wallis tests with Dunn’s correction for multiple comparisons p value; p-values resulting from pairwise tests are displayed above each structure (*** < 0.001; * < 0.05). Q2 versus canonical (p-value = 6.4×10^-32^), Q2 versus partialQ2 loss (p-value = 3.5×10^-2^), Q2 duplication versus P1 loss (p-value = 5.9×10^-98^), Q2 duplication versus Q2 loss (p-value = 8.5×10^-16^); Q2 duplication versus Q2-P1 loss (p-value = 6.2×10^-20^), canonical versus P1 loss (p-value = 2.4×10^-80^), canonical versus Q2 loss (p-value = 2.8×10^-8^), canonical versus Q2-P1 loss (p value = 1.2×10^12^), P1 loss versus Q2 loss (p-value = 2.8×10^-2^), P1 loss versus vs Q2-P1 loss (p-value = 5.6×10^-6^)

### Population distribution of other REDs

WGS cannot accurately size repeats larger than the read length (here, 150 bp), and is therefore unable to distinguish between premutation and full mutations of some REDs (e.g. *FMR1*). However, the technology can be used to determine the distribution of repeat lengths within each population, with the largest percentiles (99.9^th^ centiles) reflecting the variable presence of expanded alleles in each population. For example, the 99.9^th^ centile of repeat sizes of *DMPK* is 39 in EUR and 30 in Africans (**Table S19**, **Fig. 5A**).

Hence, we set out to extend our analysis of population differences to other RED loci. For this analysis, we used 1K GP3 data and selected RED genes that are caused by expansion of the reference sequence: *FMR1* (Fragile X syndrome), *DIP2B* (Intellectual disability FRA12A type), *ATXN8 (*SCA8), *ATXN10 (*SCA10), *LRP12* and *GIPC1* (Oculopharyngodistal myopathy 1 and 2 respectively^45^). In line with epidemiological studies, we found that for *FMR1, DIP2B* and *ATXN8* the largest percentiles are those found in Europeans, for *ATXN10* are those in Americans, and for *LRP12* are those found in East Asians. Surprisingly, we found that Africans have larger repeats in *GIPC1* compared to other ancestries (**Extended data Fig. 8, Table S21, Table S22**). The different distributions of REDs reported in this analysis reflect the relatively smaller proportion of large normal/intermediate alleles among populations, which may provide some explanation for the different frequencies of REDs in different populations.

## DISCUSSION

By analysing a cross-sectional cohort of 82,176 people, this study provides the largest population-based estimate of disease allele carrier frequency and RED distribution in different populations. We show that: i) the disease allele carrier frequency of REDs is approximately ten times higher than the previous estimates based on clinical observations, and that, based on population modelling, REDs would be predicted to affect, on average, two to three times more individuals than are currently recognised clinically; ii) while some REDs are population-specific like *JPH3* (HDL2), the majority are observed in all ancestral populations, challenging the notion that some REDs are associated with population-specific founder effects (e.g. *C9orf72*); iii) the different distribution of repeat lengths between population broadly reflects the known differences in disease epidemiology; iv) an appreciable proportion of the population carry alleles in the premutation range, and are therefore at risk of having children with REDs.

As the cohort in which we carried out this study was collected for medical sequencing purposes, we controlled for factors potentially leading to overestimating disease allele carrier frequency, such as excluding people and their relatives with neurological disorders, and checking that there was no selection bias for patients with DM1 (**Table S6**) which can cause cardiac abnormalities. Our estimates for DM1 and SCAs match those previously reported using PCR-based approaches to determine the genetic prevalence of REDs^8,19,46^, confirming the accuracy of our results.

Different factors might explain the discrepancy between the increased number of people carrying disease alleles in our cohort compared with known RED epidemiology. First, our estimates are based on large admixed cohorts, as opposed to epidemiological studies based on clinically affected individuals in smaller populations. As REDs have variable clinical presentation and age at onset, individuals with REDs may remain undiagnosed in studies in which estimates of disease frequency rely on clinical ascertainment of patients. Notably, the first descriptions of the clinical phenotype of RED were based on families collected for linkage studies with an intrinsic ascertainment bias for more severe disease manifestations, resulting in a lack of very mild cases in the phenotypic spectrum. Because of the wide spectrum of milder phenotypic presentations of REDs, the prevalence of these diseases might have been underestimated. This might be true particularly for milder forms of the disease spectrum, like DM1 for example: it is well documented that carriers of small *DMPK* expansions (50–100 repeats) have a milder disease with clinical features that may go unnoticed, especially early in their disease course^47^. In fact, we observed a large number of individuals carrying repeats in the lower end of the pathogenic range (e.g. *HTT*, *ATXN2*, and *DMPK;* **Table S9**).

On the other hand, prevalence studies are only based on individuals with manifest disease, leading to a potential bias in the disease penetrance from those who have not developed the illness. It is believed that the penetrance of REDs is characterised by a threshold effect, with people carrying an allele above a particular repeat length certainly developing the disease, as opposed to those carrying shorter repeats. Given the relationship between the size of the repeat expansion and the disease onset and progression, it is possible that individuals carrying alleles currently classified as fully penetrant (e.g. ≥40 CAG repeats in *HTT*) may sometimes remain asymptomatic. In this regard, previously published studies on HD^6^ and SBMA^48^ have suggested incomplete penetrance of repeats in the lower end of the pathogenic range.

Finally, these individuals may carry genetic modifiers of REDs, such as interspersions. We visually inspected all alleles in the pathogenic range and did not identify atypical sequence structures within the expanded repeat. Accordingly, in *HTT*, where we performed a dedicated structural analysis (**Fig. 6**), all individuals carrying a fully expanded repeat show a typical canonical structure.

The finding that a much larger number of people in the general population carry pathogenic alleles of REDs has important implications both for diagnosis and genetic counselling of RED. For diagnosis, when a patient presents with symptoms compatible with a RED, clinicians should have a higher index of suspicion of these diseases, and clinical diagnostic pathways should facilitate genetic testing for REDs. Currently, genetic testing for REDs tends to be a PCR-based targeted assay, with clinicians suspecting a RED ordering a test for a specific gene. As REDs are clinically and genetically heterogeneous with a tendency to have overlapping features, REDs may remain undiagnosed. The wider use of WGS and the advent of genetic technologies such as long-read sequencing can potentially address this by simultaneously interrogating an entire panel of RED loci^49^. The broader availability of such diagnostic tools would increase the diagnostic rate for REDs, thus closing the gap between disease incidence rates and estimates based on population genetic sequencing. As for genetic counselling, when a RED expansion is identified incidentally in an individual clinically unaffected, it would be important to address the potentially incomplete penetrance of the repeat, especially for small expansions. Further studies both in clinically affected individuals and in large clinical and genomic datasets from the general population are needed to address the full clinical spectrum and the penetrance by repeat sizes.

Our results are concordant with current epidemiological studies about the relative frequency of REDs, with the most common being DM1 and *C9orf72*-ALS/FTD (autosomal dominant) and CANVAS and sensory neuropathy (recessive). One exception is *ATXN3*, the most commonly reported SCA locus in patients affected by spinocerebellar ataxia, which was absent in our cohort. This might be due to either a recruitment bias, with individuals with overt REDs having a reduced likelihood of being recruited to such studies because of the severity of their disease (TOPMed cohort), or to the fact that there are no or very few premutation alleles (0 in 59,568 Europeans), indicating that the expansion mutation is linked to few rare ancestral haplotypes^50^, rather than being a gradual process arising from common large normal alleles like other REDs.

The presence of rare expansions of REDs previously thought to occur only in Europeans (e.g. *C9orf72)* in African and Asian populations supports diagnostic testing for them in people presenting with features of ALS-FTD independently of their ethnicity. The lowest observed rates of some REDs in some populations (for example, *FXN* and *DMPK* in East Asians), consistent with known epidemiological studies, might be due to reduced mutation rates. Further research is needed to study the potential role of population-specific *cis* and *trans* genetic modifiers of repeat expansion mutations, that underlie the marked global differences in prevalence found in the present study.

One limitation of this study is that WGS cannot accurately size repeats larger than the sequencing read length, and it is therefore not possible to accurately estimate the disease allele frequency of all RED loci. Of the 46 RED loci that have been linked to human disease^27^, we included all loci where it is technically possible to address our questions: 1) to accurately estimate the disease allele carrier frequency 16 REDs were selected; 2) to analyse the distribution of repeat lengths in different populations 6 further REDs were selected, covering a total of 22 RED loci, and providing the basis for the different prevalence of REDs in different populations. RED loci that are not included are those caused by an insertion of a non-reference sequence (currently there is no validated pipeline that can accurately size and sequence large repeat expansions in such loci, except *RFC1*), and those caused by non-pure sequences e.g. “GCN” motifs (caused by a different mutational mechanism, namely unequal allelic homologous recombination)^51^. We note that many newly discovered REDs are caused by large expansions^52^: only a broader availability of long-read sequencing technologies will facilitate addressing important questions about the frequency of these mutations.

Both 100K GP and TOPMed datasets are Euro-centric, comprising over 62% of European samples. TOPMed is more diverse, with 24% and 17% of African and American genomes respectively, which are only present at 3.2% and 2.1% frequency in the 100K GP. East and South Asian backgrounds are underrepresented in both datasets, limiting the ability to detect rarer repeat expansions in these populations. Further analyses on more heterogeneous and diverse large-scale WGS datasets are necessary not only to confirm our findings but also to shed light on additional ancestries. With regard to this, there are multiple ongoing projects with Asian populations^53^. Countries including China, Japan, Qatar, Saudi Arabia, India, Nigeria, and Turkey have all launched their own genomics projects during the last decade^54^. Analysing genomes from these programmes will yield more detail on the prevalence of REDs around the world.

Despite efforts to estimate the frequency of REDs globally and locally, there is uncertainty surrounding their true prevalence, limiting the knowledge of the burden of disease required to secure dedicated resources to support health services, such as the estimation of the numbers of individuals profiting from drug development and novel therapies, or participating in clinical trials. There are currently no disease-modifying treatments for REDs; however, both disease-specific treatments and drugs which target the mechanisms leading to repeat expansions are in development. We have established that the number of people who may benefit from such treatments is greater than previously thought.

## Acknowledgements

This research was made possible through access to data in the National Genomic Research Library, which is managed by Genomics England Limited (a wholly owned company of the Department of Health and Social Care). The National Genomic Research Library holds data provided by patients and collected by the NHS as part of their care and data collected as part of their participation in research. The National Genomic Research Library is funded by the National Institute for Health Research and NHS England. The Wellcome Trust, Cancer Research UK and the Medical Research Council have also funded research infrastructure.

This work was supported by funding from Barts charity (MGU0569) and a Medical Research Council Clinician Scientist award (MR/S006753/1) to A.T.. A.J.S. received support from NIH grants AG075051, NS105781, HD103782 and NS120241, and A.M.T. received support from NHLBI Biodata Catalyst fellowship 5120339.

Age at onset data used in the preparation of this publication for the spinocerebellar ataxia was obtained from the Rare Disease Cures Accelerator - Data and Analytics Platform (RDCA-DAP) funded by FDA Grant U18FD005320 and administered by Critical Path Institute. The data was provided to RDCA-DAP by Universitätsklinikum Bonn and Universitätsklinikum Tübingen [Universitätsklinikum Bonn and Universitätsklinikum Tübingen of datasets used by Arianna Tucci in March 2023.

Data used in the preparation of this publication for the age at onset data of myotonic dystrophy was obtained by the UK DM Patient Registry, and we would like to acknowledge the Participants and the Steering Committee.

Research reported in this paper was supported by the Office of Research Infrastructure of the National Institutes of Health under award number S10OD018522. The content is solely the responsibility of the authors and does not necessarily represent the official views of the National Institutes of Health. This work was supported in part through the computational resources and staff expertise provided by Scientific Computing at the Icahn School of Medicine at Mount Sinai.

Molecular data for the Trans-Omics in Precision Medicine (TOPMed) program was supported by the National Heart, Lung and Blood Institute (NHLBI). Genome sequencing for “NHLBI TOPMed - NHGRI CCDG: The BioMe Biobank at Mount Sinai” (phs001644.v1.p1) was performed at the McDonnell Genome Institute (3UM1HG008853-01S2). Genome sequencing for “NHLBI TOPMed: Women’s Health Initiative (WHI)” (phs001237.v2.p1) was performed at the Broad Institute Genomics Platform (HHSN268201500014C). Core support including centralized genomic read mapping and genotype calling, along with variant quality metrics and filtering were provided by the TOPMed Informatics Research Center (3R01HL-117626-02S1; contract HHSN268201800002I). Core support including phenotype harmonisation, data management, sample-identity QC, and general program coordination were provided by the TOPMed Data Coordinating Center (R01HL-120393; U01HL-120393; contract HHSN268201800001I). We gratefully acknowledge the studies and participants who provided biological samples and data for TOPMed.

The Women’s Health Initiative (WHI) program is funded by the National Heart, Lung, and Blood Institute, National Institutes of Health, U.S. Department of Health and Human Services through contracts HHSN268201600018C, HHSN268201600001C, HHSN268201600002C, HHSN268201600003C, and HHSN268201600004C. This manuscript was not prepared in collaboration with investigators of the WHI, and does not necessarily reflect the opinions or views of the WHI investigators, or NHLBI.

The Atherosclerosis Risk in Communities study has been funded in whole or in part with Federal funds from the National Heart, Lung, and Blood Institute, National Institute of Health, Department of Health and Human Services, under contract numbers (HHSN268201700001I, HHSN268201700002I, HHSN268201700003I, HHSN268201700004I, and HHSN268201700005I). The authors thank the staff and participants of the ARIC study for their important contributions MESA and the MESA SHARe project are conducted and supported by the National Heart, Lung, and Blood Institute (NHLBI) in collaboration with MESA investigators. Support for MESA is provided by contracts HHSN268201500003I, N01-HC-95159, N01-HC-95160, N01-HC-95161, N01-HC-95162, N01-HC-95163, N01-HC95164, N01-HC-95165, N01-HC-95166, N01-HC-95167, N01-HC-95168, N01-HC-95169, UL1-TR-001079, UL1-TR000040, UL1-TR-001420, UL1-TR-001881, and DK063491.

The Jackson Heart Study (JHS) is supported and conducted in collaboration with Jackson State University (HHSN268201800013I), Tougaloo College (HHSN268201800014I), the Mississippi State Department of Health (HHSN268201800015I/HHSN26800001) and the University of Mississippi Medical Center (HHSN268201800010I, HHSN268201800011I and HHSN268201800012I) contracts from the National Heart, Lung, and Blood Institute (NHLBI) and the National Institute for Minority Health and Health Disparities (NIMHD). The authors also wish to thank the staff and participants of the JHS.

This research was supported by contracts HHSN268201200036C, HHSN268200800007C, HHSN268201800001C, N01HC55222, N01HC85079, N01HC85080, N01HC85081, N01HC85082, N01HC85083, N01HC85086, and grants U01HL080295 and U01HL130114 from the National Heart, Lung, and Blood Institute (NHLBI), with additional contribution from the National Institute of Neurological Disorders and Stroke (NINDS). Additional support was provided by R01AG023629 from the National Institute on Aging (NIA). A full list of principal CHS investigators and institutions can be found at CHS-NHLBI.org.

This research used data generated by the COPDGene study, which was supported by NIH grants U01 HL089856 and U01 HL089897. The COPDGene project is also supported by the COPD Foundation through contributions made by an Industry Advisory Board composed of Pfizer, AstraZeneca, Boehringer Ingelheim, Novartis, and Sunovion.

The Framingham Heart Study is conducted and supported by the National Heart, Lung, and Blood Institute (NHLBI) in collaboration with Boston University (Contract No. N01-HC-25195, HHSN268201500001I and 75N92019D00031). This manuscript was not prepared in collaboration with investigators of the Framingham Heart Study and does not necessarily reflect the opinions or views of the Framingham Heart Study, Boston University, or NHLBI.

The Mount Sinai BioMe Biobank is supported by The Andrea and Charles Bronfman Philanthropies.

This research utilised Queen Mary’s Apocrita HPC facility, supported by QMUL Research-IT^55^.

## Authors’ contribution statement

K.I. and A.T. conceptualised the research project. K.I., B.J. M.Z., D.G. C.C., S.F., P.G, L.M., M.S., V.E.P., A.T. A.J.S, conducted data analysis, interpreted statistical findings and created visual representations of the data. K.I., B.J., M.Z., P.G., A.M.T., S.J.G., V.G.D., D.J.H.M, C.R., A.T. performed quality check visually inspecting pileup plots corresponding to repeat expansions. J.V. analysed the carrier frequency for RFC1 within the 100K GP. C.M.B., H.W., C.T., S.T., J.D.L and D.R.L. provided data from HD and DM1 patient registries. A.C. provided valuable insights into the genetics of RFC1. Funding and supervision: A.T., A.J.S., P.F., M.J.C., J.H, H.H. Writing—original draft: K.I., A.T., A.J.S., A.D., D.J.H.M., M.Z., C.R., and M.S. meticulously reviewed and edited the manuscript for clarity, accuracy, and coherence. All authors carefully reviewed the manuscript, offering pertinent feedback that enhanced the study’s quality, and ultimately approved the final version.

## Methods

### Ethics Statement Inclusion & Ethics

The 100 000 Genomes Project is a UK programme to assess the value of whole genome sequencing in patients with unmet diagnostic needs in rare disease and cancer. Following ethical approval for the 100 000 Genomes Project by the East of England Cambridge South Research Ethics Committee (reference 14/EE/1112), including for data analysis and return of diagnostic findings to the patients, these patients were recruited by health-care professionals and researchers from 13 Genomic Medicine Centres in England, and were enrolled in the project if they or their guardian provided written consent for their samples and data to be used in research, including this study.

For ethics statements for the contributing TOPMed studies, full details are provided in the original description of the cohorts^1^.

### Whole genome sequencing datasets

Both 100,000 Genomes Project (100K GP) and Trans-Omics for Precision Medicine (TOPMed) include whole genome sequencing (WGS) data optimal to genotype short DNA repeats: WGS libraries generated using PCR-free protocols, sequenced at 150 base-pair read-length, and with a 35x mean average coverage (Table S1).

For both the 100K GP and TOPMed cohorts, the following genomes were selected: i) WGS from genetically unrelated individuals (see Ancestry and relatedness inference below); ii) WGS from people not presenting with a neurological disorder - these people were excluded to avoid overestimating the frequency of a repeat expansion due to individuals recruited due to symptoms related to a RED.

The TOPMed project has generated omics data, including WGS, on over 180,000 individuals with heart, lung, blood, and sleep disorders (see <u>NHLBI Trans-Omics for Precision Medicine WGS-About TOPMed</u> (nih.gov)). TOPMed has incorporated samples gathered from dozens of different cohorts, each collected using different ascertainment criteria. The specific TOPMed cohorts included in this study are described in **Table S23**.

To analyse the distribution of repeat lengths of RED genes for which EH is not able to discriminate between the premutation/reduced penetrance and full mutation alleles in different populations, we used the 1000 Genomes Project phase 3 (1K GP3) as the WGS data are more equally distributed across the continental groups (**Table S2**). Genome sequences with read lengths of ∼150bp were considered, with an average minimum depth of 30x (**Table S1**).

### Ancestry and relatedness inference

For relatedness inference WGS VCFs were aggregated with Illumina’s agg or gvcfgenotyper (https://github.com/Illumina/gvcfgenotyper). All genomes passed the following quality control (QC) criteria: cross-contamination <5% (VerifyBamId)^2^, mapping rate >75%, mean-sample coverage >20, and insert size > 250 bp. No variant QC filters were applied in the aggregated dataset, but the VCF filter was set to ‘PASS’ for variants which passed GQ (genotype quality), DP (depth), missingness, allelic imbalance, and Mendelian error filters. From here, by using a set of ∼65,000 high-quality SNPs, a pairwise kinship matrix was generated using the PLINK2 implementation of the KING-Robust algorithm (www.cog-genomics.org/plink/2.0/)^3^. For relatedness, PLINK2 ‘--king-cutoff’ (www.cog-genomics.org/plink/2.0/) relationship-pruning algorithm ^3^ was used with a threshold of 0.044. These were then partitioned into ‘related’ (up to, and including, 3^rd^-degree relationships) and ‘unrelated’ sample lists. Only unrelated samples were selected for this study.

1K GP3 data was used to infer ancestry, by taking the unrelated samples and calculating the first 20 PCs using GCTA2. We then projected the aggregated data (100K GP and TOPMed separately) onto 1K GP3 PC loadings, and a random forest model was trained to predict ancestries based on 1) First 8 1K GP3 PCs, 2) setting ‘Ntrees’ to 400, and 3) train and predict on 1kPG3 five broad super-populations: African (AFR), Admixed American (AMR), East Asian (EAS), European (EUR), and South Asian (SAS).

In total, the following WGS data were analysed: 34,190 individuals in the 100K GP; 47,986 in TOPMed; 2,504 in the 1K GP3. The demographics describing each cohort can be found in Table S2.

### Correlation between PCR and ExpansionHunter

Results were obtained on samples tested as part of routine clinical assessment from patients recruited to the 100K GP. Repeat expansions were assessed by polymerase chain reaction (PCR) amplification and fragment analysis. Southern blotting was performed for large *C9orf72* and *NOTCH2NL* expansions as previously described^4^.

A dataset was set up from the 100KGP samples comprising a total of 681 genetic tests with PCR-quantified lengths across 15 loci: AR, ATN1, ATXN1, ATXN2, ATXN3, ATXN7, CACNA1A, DMPK, C9orf72, FMR1, FXN, HTT, NOTCH2NL, PPP2R2B, TBP (Table S3).

Overall, this dataset comprised PCR and correspondent EH estimates from a total of 1,291alleles: 1,146 normal, 44 premutation and 101 full mutation. Fig. S3A shows the swim lane plot of EH repeat sizes after visual inspection classified as normal (blue), premutation or reduced penetrance (yellow) and full mutation (red). This data shows that EH correctly classifies 28/29 premutations and 85/86 full mutations for all loci assessed, after excluding *FMR1* (Table S3; Table S4). For this reason, this locus has not been analysed to estimate the premutation and full mutation alleles carrier frequency. The two alleles with a mismatch are changes of one repeat unit in *TBP* and *ATXN3*, changing the classification (Table S3). Fig. S3B shows the distribution of repeat sizes quantified by PCR compared to those estimated by EH after visual inspection, split by super-population. Pearson correlation (R) calculated separately for alleles larger (for Europeans, n=864) and shorter (n=76) than the read length (i.e. 150bp).

### Repeat expansion genotyping and visualisation

ExpansionHunter v3.2.2 (EH) software package was used for genotyping repeats in disease-associated loci^5,6^. EH assembles sequencing reads across a predefined set of DNA repeats using both mapped and unmapped reads (with the repetitive sequence of interest) to estimate the size of both alleles from an individual.

REViewer software package was used to enable direct visualisation of haplotypes and corresponding read pileup of the EH genotypes^7^. Table S24 includes the genomic coordinates for the loci analysed. Table S5 lists repeats before and after visual inspection. Pileup plots are available upon request.

### Computation of genetic prevalence

The frequency of each repeat size across the 100K GP and TOPMed genomic datasets was determined. Genetic prevalence was calculated as the number of genomes with repeats exceeding the premutation and full mutation cut-offs (**Table 1**) for autosomal dominant and X-linked REDs (**Table S7**); for autosomal recessive REDs, the total number of genomes with monoallelic or biallelic expansions was calculated, compared to the overall cohort **(Table S8)**.

Overall unrelated and non-neurological disease genomes corresponding to both programmes were considered, breaking down by ancestry.

CARRIER FREQUENCY ESTIMATE (1 in x)

- freq_carrier = round(total_unrel / total_exp_after_VI_locus, digits = 0) where

- ‘total_unrel’ is the total number of unrelated genomes

- ‘total_exp_after_VI_locus’ is the total number of genomes that have a repeat expansion beyond premutation or full-mutation after visual inspection (per each locus)

CONFIDENCE INTERVALS:

- n = total number of unrelated genomes

- p = total expansions / total number of unrelated genomes

- q = 1 - p

- z = 1.96

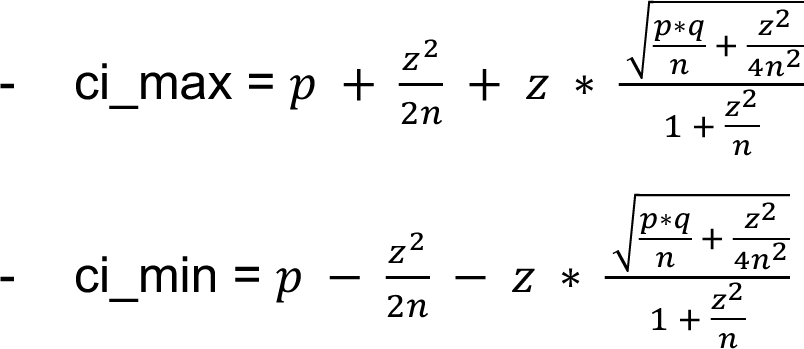

PREVALENCE ESTIMATE (x in 100,000)

x = 100,000/freq_carrier new_low_ci = 100000*ci_max_final new_high_ci = 100000*ci_min_final

Modelling disease prevalence using carrier frequency

The total number of expected people with the disease caused by the repeat expansion mutation in the population (𝑀) was estimated as:

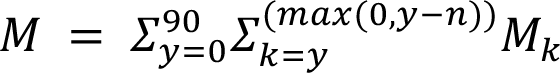

where 𝑀, is the expected number of new cases at age 𝑘 with the mutation and 𝑛 is survival length with the disease in years.

𝑀, is estimated as 𝑀, = 𝑓 ∗ 𝑁, ∗ 𝑝, where 𝑓 is the frequency of the mutation, 𝑁, is the number of people in the population at age 𝑘 (according to Office of National Statistics, ONS^8^), and 𝑝, is the proportion of people with the disease at age 𝑘, estimated at the number of the new cases at age 𝑘 (according to cohort studies and international registries) divided by the total number of cases.

To estimate the expected number of new cases by age group, the age at onset distribution of the specific disease, available from cohort studies or international registries, was used. For *C9orf72*-disease, we tabulated the distribution of disease onset of 811 patients with *C9orf72*-ALS pure and overlap FTD, and 323 patients with *C9orf72*-FTD pure and overlap ALS^22^. HD onset was modelled using data derived from a cohort of 2,913 individuals with HD described by Langbehn et al.^23^; and DM1 was modelled on a cohort of 264 non-congenital patients derived from the UK Myotonic Dystrophy patient registry^24^. Data from 157 patients with SCA2 and *ATXN2* allele size equal to or higher than 35 repeats from EUROSCA were used to model the prevalence of SCA2^25^. From the same registry, data from 91 patients with SCA1 and *ATXN1* allele sizes equal to or higher than 44 repeats and of 107 patients with SCA6 and *CACNA1A* allele sizes equal to or higher than 20 repeats were used to model disease prevalence of SCA1 and SCA6, respectively.

As some REDs have reduced age-related penetrance, e.g. *C9orf72*-carriers may not develop symptoms even after 90 years of age^22^, age-related penetrance was obtained as follows: as regards *C9orf72*-ALS/FTD, it was derived from the red curve in **Fig.2** (data available at https://github.com/nam10/C9_Penetrance) reported by Murphy et al^22^ and was used to correct *C9orf72*-ALS and *C9orf72*-FTD prevalence by age. For HD, age-related penetrance for a 40 CAG repeat carrier was provided by DRL, based on his work^23^.

DETAILED DESCRIPTION OF THE METHOD THAT EXPLAINS TABLE S10-S16:

Both the general UK population and age at onset distribution of each disease were tabulated (**Tables S10-S16**, column B and column C). After standardisation over the total number (**Tables S10-S16**, column D), the onset count was multiplied by the carrier frequency of the genetic defect (**Tables S10-S16**, column E) and then multiplied by the corresponding general population count for each age group, to obtain the estimated number of people in the UK developing each specific disease by age group (**Tables S10-S11**, column G, **Tables S12-S16** column F). This estimate was further corrected by the age-related penetrance of the genetic defect where available (e.g., *C9orf72*-ALS and FTD) (**Tables S10-S11**, columns F). Finally, to account for disease survival, we performed a cumulative distribution of prevalence estimates grouped by a number of years equal to the median survival length for that disease (**Tables S10-S11**, column H, **Tables S12-S16** column G). The median survival length (𝑛) used for this analysis is: 3 years for *C9orf72*-ALS^9^, 10 years for *C9orf72*-FTD^9^, 15 years for HD^14^ (40 CAG repeat carriers), 15 years for SCA2 and for SCA1^18^; for SCA6 normal life expectancy was assumed. For DM1, since life expectancy is partly related to the age of onset, the mean age of death was assumed to be 45 years for patients with childhood onset and 52 years for early adult-onset patients (10-30 years)^26^, while no age of death was set for DM1 patients with onset after 31 years. Since survival is approximately 80% after 10 years^20^, we subtracted 20% of the predicted affected individuals after the first 10 years. Then, survival was assumed to proportionally decrease in the following years until the mean age of death for each age group was reached.

The resulting estimated prevalences of *C9orf72*-ALS/FTD, HD, SCA2, DM1, SCA1, and SCA6 by age group were plotted in Fig.3 (dark blue area). The literature reported prevalence by age for each disease was obtained by dividing the new estimated prevalence by age by the ratio between the two prevalences, and represented as a light blue area.

To compare the new estimated prevalence to the clinical disease prevalence reported in the literature for each disease, we employed figures calculated in European populations, as it is closer to the UK population in terms of ethnic distribution:

i) *C9orf72*-FTD: the median prevalence of FTD was obtained from studies included in the systematic review by Hogan and colleagues^12^ (83.5 in 100,000). Since 4-29% of FTD patients carry a *C9orf72* repeat expansion^13^, we calculated *C9orf72*-FTD prevalence by multiplying this proportion range by median FTD prevalence (3.3 - 24.2 in 100,000, mean 13.78 in 100,000); ii) *C9orf72*-ALS: The reported prevalence of ALS is 5-12 in 100,000^10^ and *C9orf72* repeat expansion is found in 30%-50% of individuals with familial forms and in 4%-10% of people with sporadic disease^11^. Given that ALS is familial in 10% of cases and sporadic in 90%, we estimated the prevalence of *C9orf72*-ALS by calculating the [(0.4 of 0.1) + (0.07 of 0.9)] of known ALS prevalence of 0.5-1.2 in 100,000 (mean prevalence is 0.8 in 100,000); iii) HD prevalence ranges from 0.4 in 100,000 in Asian countries^15^ to 10 in 100,000 in Europeans^16^, and the mean prevalence is 5.2 in 100,000. 40-CAG repeat carriers represent 7.4% of patients clinically affected by HD according to the Enroll-HD^17^ version 6. Considering an average reported prevalence of 9.7 in 100,000 Europeans, we calculated a prevalence of 0.72 in 100,000 for symptomatic 40-CAG carriers; iv) DM1 is much more frequent in Europe than in other continents, with figures of 1 in 100,000 in some areas of Japan^27^. A recent meta-analysis has found an overall prevalence of 12.25 per 100,000 individuals in Europe, which we used in our analysis^21^.

Given that the epidemiology of autosomal dominant ataxias varies among countries^19^ and no precise prevalence figures derived from clinical observation are available in the literature, we approximated SCA2, SCA1, and SCA6 prevalence figures to be equal to 1 in 100,000.

### Local ancestry prediction

#### 100K GP

For each RE locus and for each sample with a premutation or a full mutation, we obtained a prediction for the local ancestry in a region of +/-5Mb around the repeat, as follows:

1. We extracted VCF files with SNPs from the selected regions and phased them with SHAPEIT v4. As a reference haplotype set, we used non-admixed individuals from the 1kG project. Additional non-default parameters for SHAPEIT: --mcmc-iterations 10b,1p,1b,1p,1b,1p,1b,1p,10m –pbwt-depth 8.

2. The phased VCFs were merged with non-phased genotype prediction for the repeat length, as provided by ExpansionHunter. These combined VCFs were then phased again using Beagle v4.0. This separate step is necessary because SHAPEIT does not accept genotypes with more than the two possible alleles (as is the case for repeat expansions which are polymorphic).

3. Finally, we attributed local ancestries to each haplotype with RFmix, using the global ancestries of the 1kG samples as a reference. Additional parameters for RFmix: -n 5-G 15 -c 0.9 -s 0.9 –reanalyze-reference

#### TOPMed

The same method was followed for TOPMed samples, except that in this case the reference panel also included individuals from the Human Genome Diversity Project.

1, We extracted SNPs with maf >= 0.01 that were within +/-5 Mb of the tandem repeats and ran beagle (version 5.4, beagle.22Jul22.46e) on these SNPs to perform phasing with parameters burnin=10 and iterations=10.

#### SNP phasing using beagle

java -jar ./beagle.22Jul22.46e.jar \ gt=${input} \

ref=../RefVCF/hgdp.tgp.gwaspy.merged.chr${chr}.merged.cleaned.vcf.gz \ out=Topmed.SNPs.maf0.001.chr${prefix}.beagle \

chrom=$region \ burnin=10 \ iterations=10 \

map=./genetic_maps/plink.chr${chr}.GRCh38.map \ nthreads=${threads} \

impute=false

2. Next, we merged the unphased tandem repeat genotypes with the respective phased SNP genotypes using the bcftools. We used beagle version r1399, incorporating the parameters burnin-its=10, phase-its=10, and usephase=true. This version of beagle allows multiallelic Tander Repeat to be phased with SNPs.

java -jar ./beagle.r1399.jar \ gt=${input} \

out=${prefix} \ burnin-its=10 \ phase-its=10 \

map=./genetic_maps/plink.${chr}.GRCh38.map \ nthreads=${threads} \

usephase=true

3. To conduct local ancestry analysis (LAI), we used RFMIX^28^ with the parameters -n 5 -e 1 -c 0.9 -s 0.9 and -G 15. We utilised phased genotypes of 1,000 genomes as a reference panel^29^.

time rfmix \

-f $input \

-r ../RefVCF/hgdp.tgp.gwaspy.merged.${chr}.merged.cleaned.vcf.gz \

-m samples_pop \

-g genetic_map_hg38_withX_formatted.txt \

--chromosome=$c \

-n 5 \

-e 1 \

-c 0.9 \

-s 0.9 \

-G 15 \

--n-threads=48 \

-o $prefix

### Distribution of repeat lengths in different populations

#### Repeat size distribution analysis

The distribution of each of the 16 RE loci where our pipeline enabled discrimination between the premutation/reduced penetrance and the full mutation was analysed across the 100K GP and TOPMed datasets (Fig. 5A, Fig. S6), while the distribution of larger repeat expansions was analysed in the 1K GP3 (Fig. 7). For each gene, the distribution of the repeat size across each ancestry subset was visualised as a density plot and as a box-blot; moreover, the 99.9^th^ percentile and the threshold for intermediate and pathogenic ranges were highlighted (Table S19, Table S21, Table S22).

#### Correlation between intermediate and pathogenic repeat frequency

The percentage of alleles in the intermediate and in the pathogenic range (premutation plus full mutation) was computed for each population (combining data from 100K GP with TOPMed) for genes with a pathogenic threshold below or equal to 150 bp. The intermediate range was defined as either the current threshold reported in the literature^30–34^ (*ATXN1*=36; *ATXN2*=31; *ATXN7*=28; *CACNA1A*=18; *HTT*=27) or as the reduced penetrance/premutation range according to **Table 1** for those genes where the intermediate cutoff is not defined (*AR, ATN1, DMPK, JPH3*, and *TBP*) (**Table S20**). Genes where either the intermediate or pathogenic alleles were absent across all populations were excluded. Per population, intermediate and pathogenic allele frequencies (percentages) were displayed as a scatter plot using R and the package tidyverse, and correlation was assessed using Spearman’s rank correlation coefficient with the package ggpubr and the function stat_cor (**Fig. 5B, Fig. S7**).

#### *HTT* structural variation analysis

We developed an in-house analysis pipeline named Repeat Crawler (RC) to ascertain the variation in repeat structure within and bordering the *HTT* locus. Briefly, RC takes the mapped BAMlet files from ExpansionHunter as input and outputs the size of each of the repeat elements in the order that is specified as input to the software (i.e. Q1, Q2 and P1). To ensure that the reads that RC analyses are reliable, we restrict our analysis to only utilise spanning reads. In order to haplotype the CAG repeat size to its corresponding repeat structure, RC utilised only spanning reads that encompassed all the repeat elements including the CAG repeat (Q1). For larger alleles that could not be captured by spanning reads, we re-ran RC excluding Q1. For each individual, the smaller allele can be phased to its repeat structure using the first run of RC and the larger CAG repeat is phased to the second repeat structure called by RC in the second run. RC is available at https://github.com/chrisclarkson/gel/tree/main/HTT_work.

To characterise the sequence of the *HTT* structure, we used 66,838 alleles from 100K GP genomes. These correspond to 97% of the alleles, with the remaining 3% consisting of calls where EH and RC did not agree on either the smaller or bigger allele.

## Data availability

For the 100K GP, full data is available in the Genomic England Secure Research Environment. Access is controlled to protect the privacy and confidentiality of participants in the Genomics England 100,000 Genomes Project and to comply with the consent given by participants for use of their healthcare and genomic data. Access to full data is permitted through the Research Network (https://www.genomicsengland.co.uk/research/academic/join-research-network).

For TOPMed, a detailed description of the TOPMed participant consents and data access is provided in Box 1^1^. TOPMed data used in this manuscript are available through dbGaP. The dbGaP accession numbers for all TOPMed studies referenced in this paper are listed in Extended Data Tables 23^1^. A complete list of TOPMed genetic variants with summary level information used in this manuscript is available through the BRAVO variant browser (<u>bravo.sph.umich.edu</u>). The TOPMed imputation reference panel described in this manuscript can be used freely for imputation through the NHLBI BioData Catalyst at the TOPMed Imputation Server (https://imputation.biodatacatalyst.nhlbi.nih.gov/). DNA sequence and reference placement of assembled insertions are available in VCF format (without individual genotypes) on dbGaP under the TOPMed GSR accession <u>phs001974</u>.

For the 1000 Genomes Project, the WGS datasets are available from the European Nucleotide Archive under accessions PRJEB31736 (unrelated samples) and PRJEB36890 (related samples).

## Code availability

The following GitHub repositories used in this work are free to access:

- Expansion Hunter v3.2.2 (to estimate the repeat size within defined loci): https://github.com/Illumina/ExpansionHunter

- REViewer v0.2.7 (to generate pileup plots for quality check): https://github.com/Illumina/REViewer

- ExpansionHunter_Classifier (March 2024 release, to automatically run quality assessment of EHv322 call): https://github.com/bharatij/ExpansionHunter_Classifier

- Code to analyse repeat structure across *HTT*: https://github.com/chrisclarkson/gel/tree/main/HTT_work

- gvcfgenotyper (to merge gVCF files, when inferring ancestry across genomes within the 100K GP and TOPMed datasets): https://github.com/Illumina/gvcfgenotyper

- To compute survival curve analysis for *C9orf72*: https://github.com/nam10/C9_Penetrance

