## extended data figure legend for "Increased frequency of repeat expansion mutations across different populations"

| **Extended Figure #** | **Figure title** | **Figure Legend** |
| --- | --- | --- |
| Extended Data Fig. 1 | Population pyramid of (A) the 100K GP and (B) TOPMed cohorts. |  |
| Extended Data Fig. 2 | Principal components of genetic ancestry | First two principal components derived from PCA on A) the 100K GP and B) TOPMed samples respectively |
| Extended Data Fig. 3 | Experimental estimations of repeat sizes using PCR versus genotypes generated by ExpansionHunter v3.2.2. | A). Swim lane plot showing sizes of repeat expansions predicted by ExpansionHunter across 681 samples with expansion calls. Each genome is represented by two points, one corresponding to each allele for each locus, except for those on the X chromosome (i.e. *FMR1* and *AR*) in males, for which only one point is shown. Points indicate the repeat length estimated by ExpansionHunter after visual inspection and the colours indicate the repeat size as assessed by PCR (blue represents non-expanded; red represents expanded). The regions are shaded to indicate non-expanded (blue), premutation (yellow), and expanded (red) ranges for each gene, as indicated in Table 1. Blue points in yellow or red-shaded regions indicate false positives and red points in blue-shaded regions indicate false negatives. The individual calls are provided in Table S3. B). Points indicate the RE size estimated by both PCR and EH v3.2.2 split by super-population. We show the R correlation coefficient calculated using Pearson’s equation and two-tailed P values. Exact p-values for the regression model: AFR (1.1x10^-28^), AMR (2.1x10^-29^), EUR (1.7x10^-168^), and SAS (1.3x10^–80^). |
| Extended Data Fig. 4 | Distribution of repeat size alleles within the combined 100K GP and TOPMed cohort. | Allele frequency (percentage) predicted by ExpansionHunter in the combined 100K GP and TOPMed cohorts. The regions are shaded to indicate non-expanded (blue), premutation (yellow), and full mutation expanded (red) ranges for each gene, as indicated in Table 1. For *RFC1,* repeat sizes beyond 30 are shaded as repeat sizes beyond this threshold may represent expanded alleles |
| Extended Data Fig. 5 | PC values of genomes carrying normal and pathogenic alleles | Principal component (PC) values on all genomes within (A) the 100K GP and (B) TOPMed cohorts. Black dots represent genomes having a repeat size beyond premutation and full mutation range for X-linked and autosomal dominant loci, split by locus. For recessive loci, the plot shows genomes carrying monoallelic and biallelic expansions. Note that *RFC1* has only been analysed in the 100K GP dataset due to code availability. Note that *ATXN3* is missing from the 100K GP panels as there are no pathogenic alleles in this cohort (**Table S7**). |
| Extended Data Fig. 6 | Distribution of repeat size alleles in different populations in the combined cohort (100KP and TOPMed) | Half-violin plots showing the distribution of alleles in different populations for 6 loci excluded from the correlation analysis from the combined 100K GP and TOPMed cohort (African = 12,786;  American = 5,674; East Asian = 1,266; European = 59,568; South Asian = 2,882). Boxplots highlight the interquartile range and median, and black dots show values outside 1.5 times interquartile ranges. Red dots mark the 99.9^th^ percentile for each population and locus. Vertical bars indicate the intermediate and pathogenic allele thresholds (**Table S20**). |
| Extended Data Fig. 7 | Frequency of intermediate alleles versus frequency of pathogenic alleles by population | The scatter plots show the frequency of intermediate allele carriers (x-axis) against the frequency of pathogenic allele carriers (y-axis), based on the thresholds in Table S20, split by population. Data points are divided by gene (n=10), and size represents the total number of intermediate alleles. Correlations were computed using the Spearman method. |
| Extended Data Fig. 8 | Distribution of repeat size alleles by population in the 1K GP | Distribution of disease RE sizes for 22 genes within the 1K GP3 split by population (African = 661; American = 347; East Asian = 504; European = 503; South Asian = 489). Half-violin plots show the distribution of alleles, while boxplots highlight the interquartile range and median, and black dots show values outside 1.5 times interquartile ranges. Red dots mark the 99.9th percentile for each population and locus. Repeat size mean;median (Q1-Q3) among all ancestries are in Table S19. |
| Choose an item. |  |  |
| Choose an item. |  |  |
