## Supplementary figures and images for "Increased frequency of repeat expansion mutations across different populations"

### Extended data figure 1

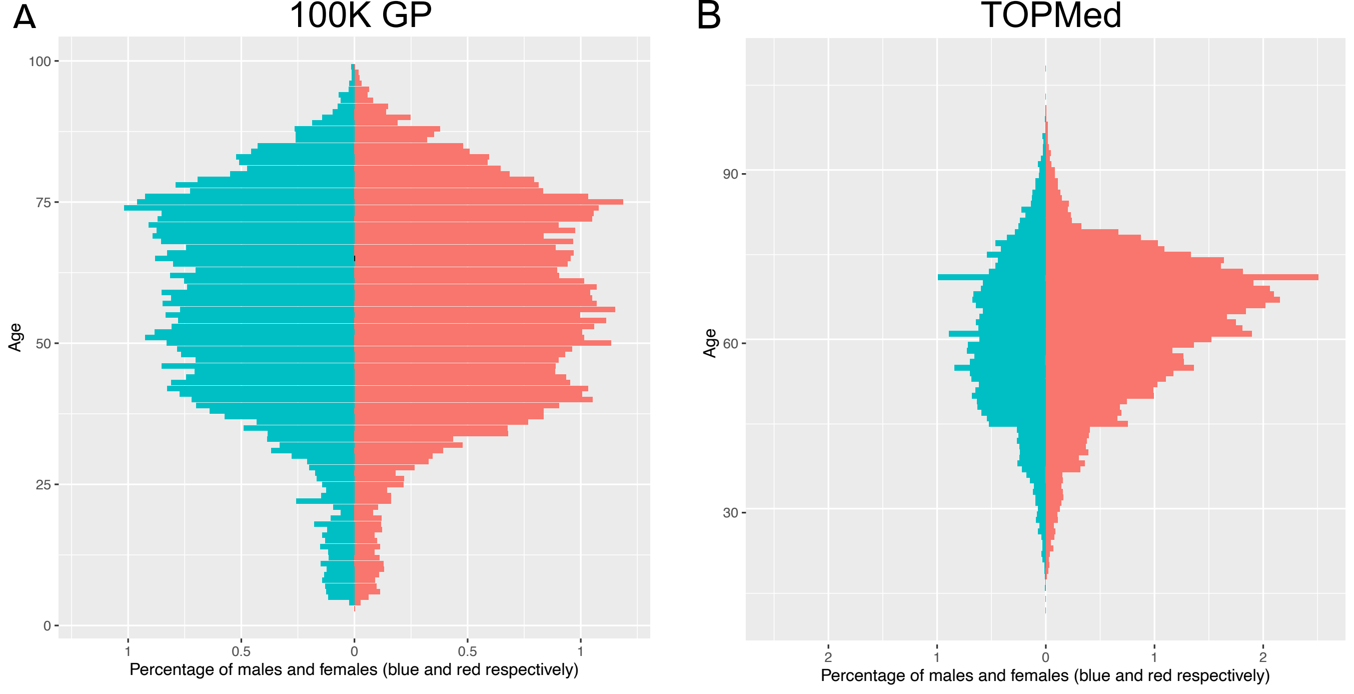

### Extended data figure 2

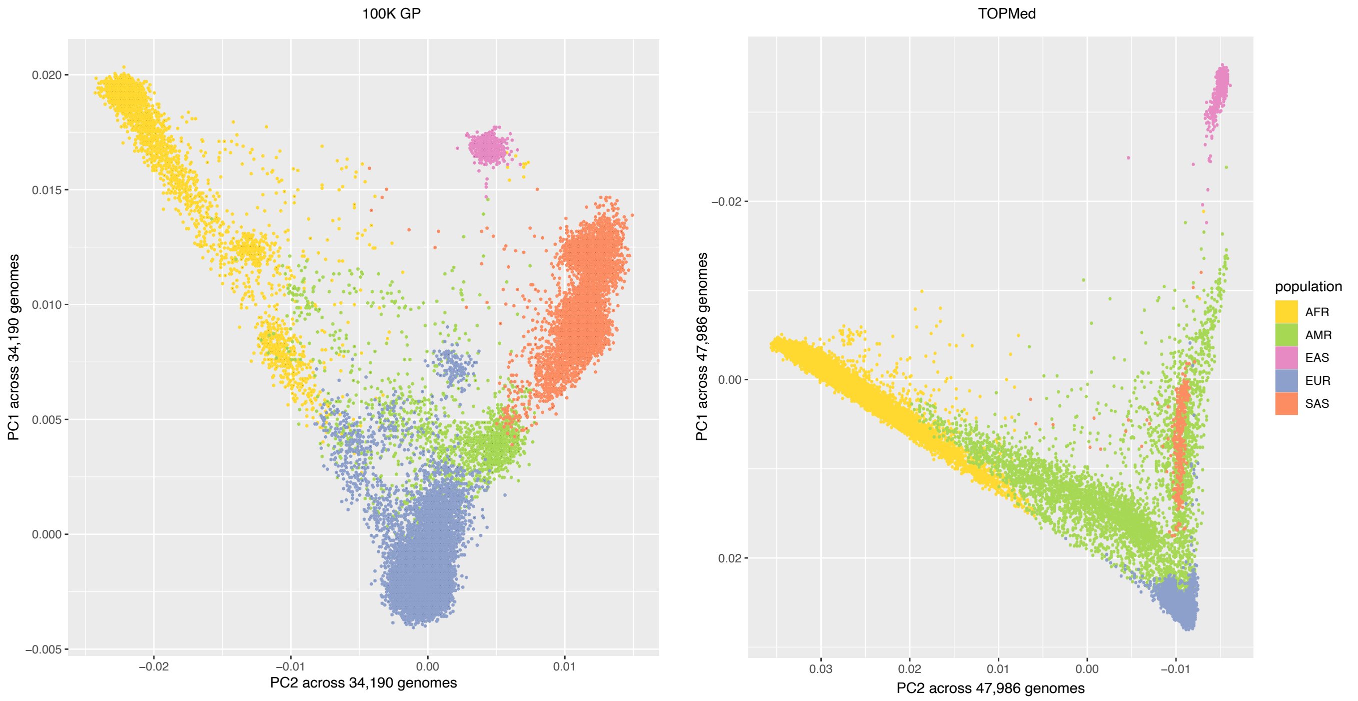

### Extended data figure 3

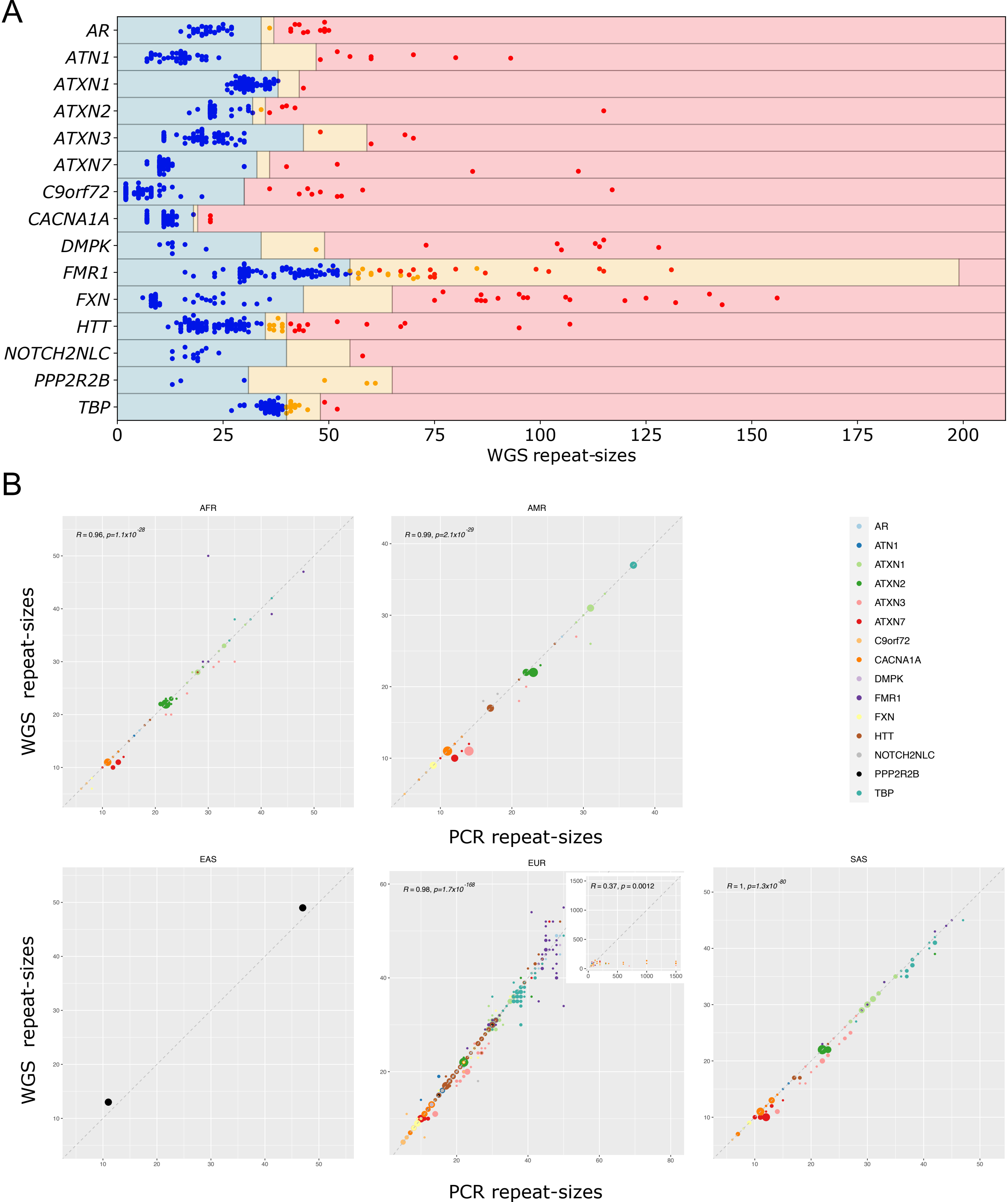

### Extended data figure 4

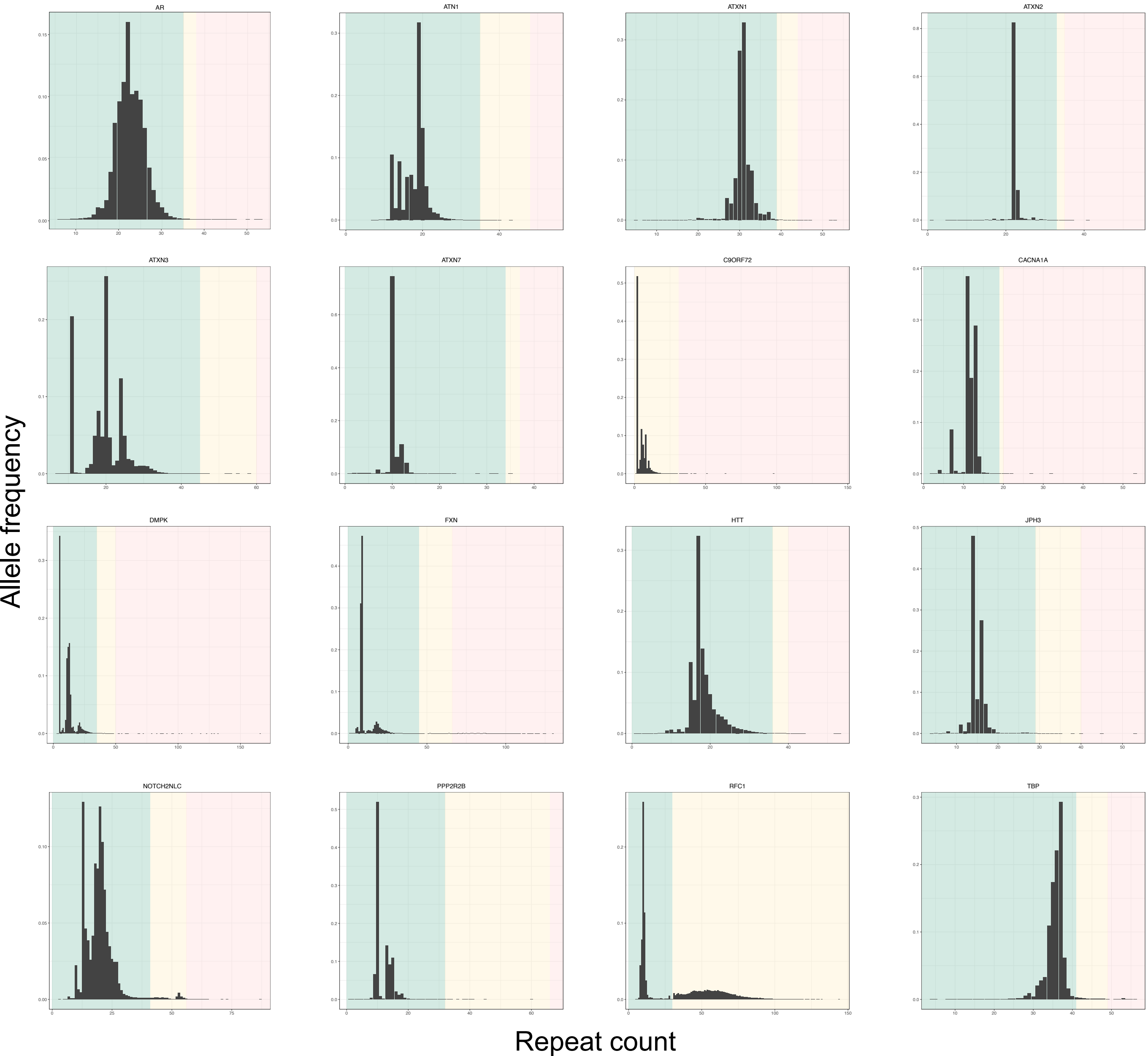

### Extended data figure 5

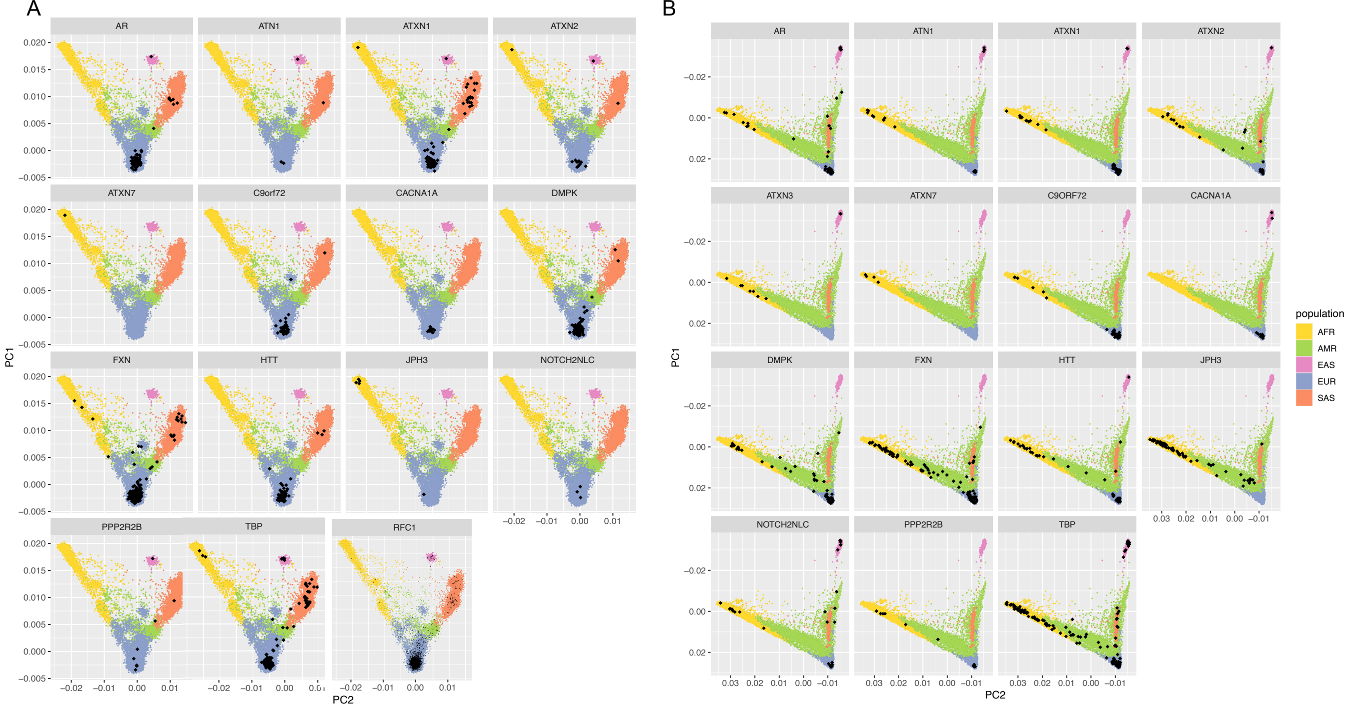

### Extended data figure 6

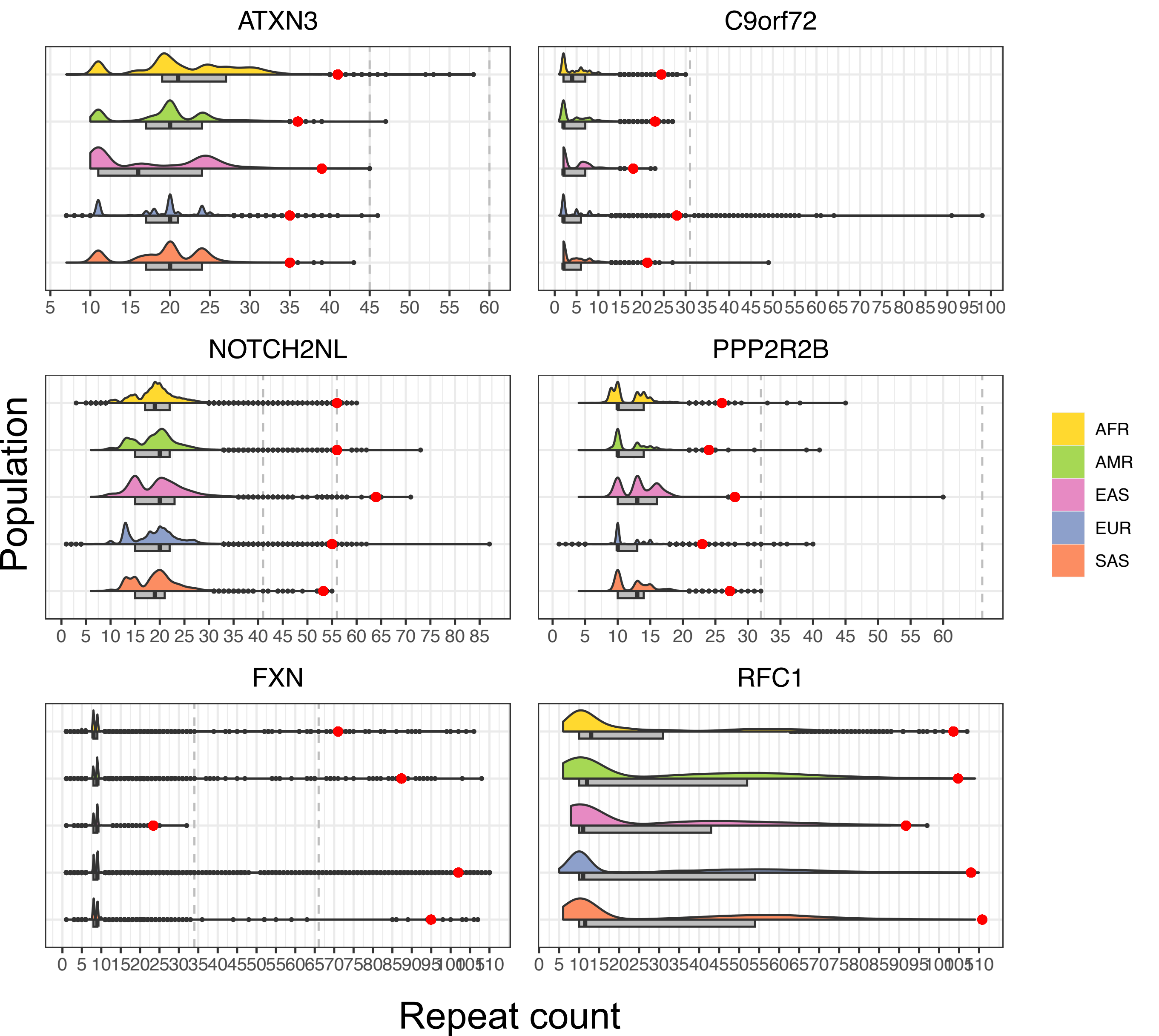

### Extended data figure 7

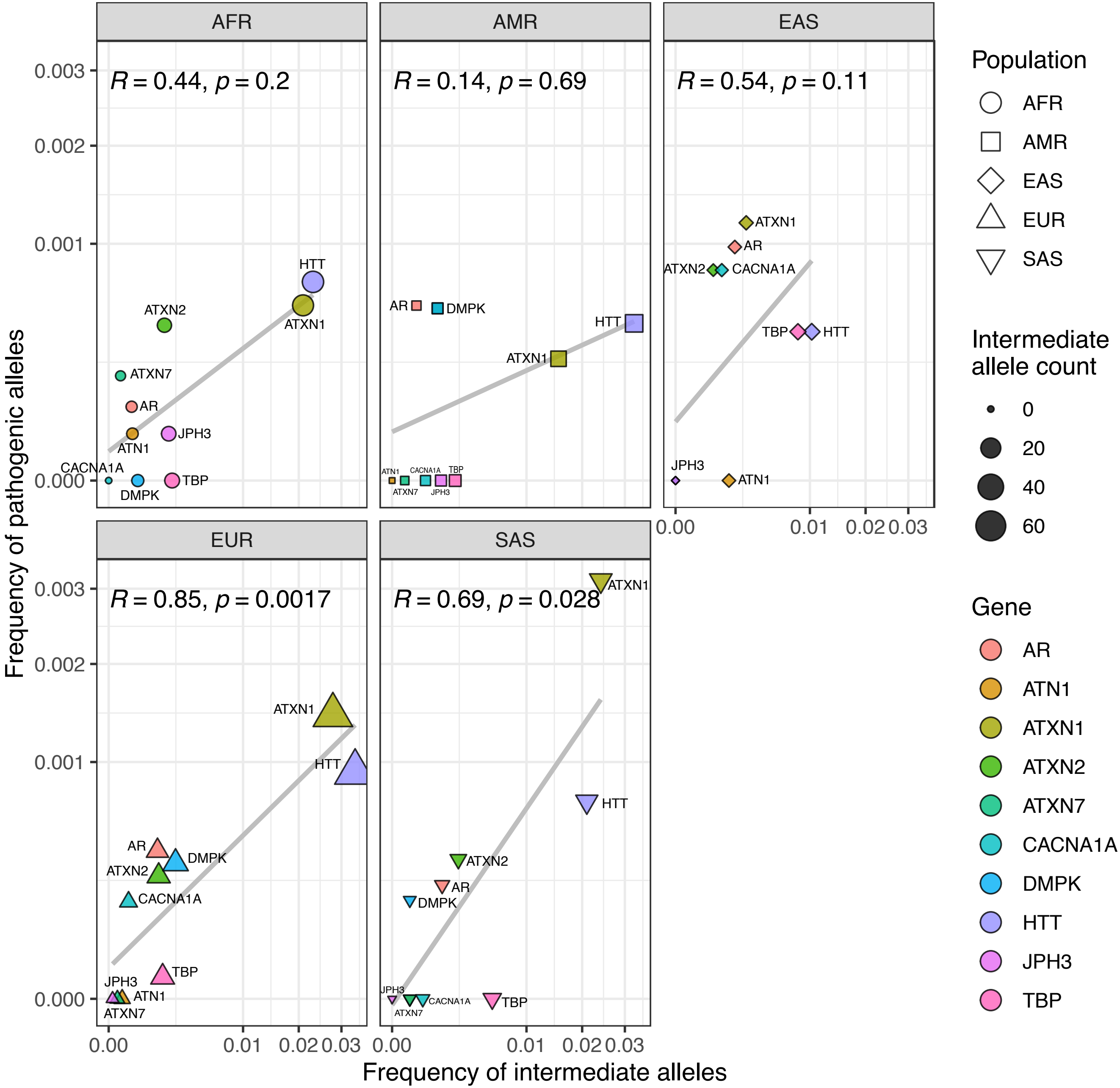

### Extended data figure 8

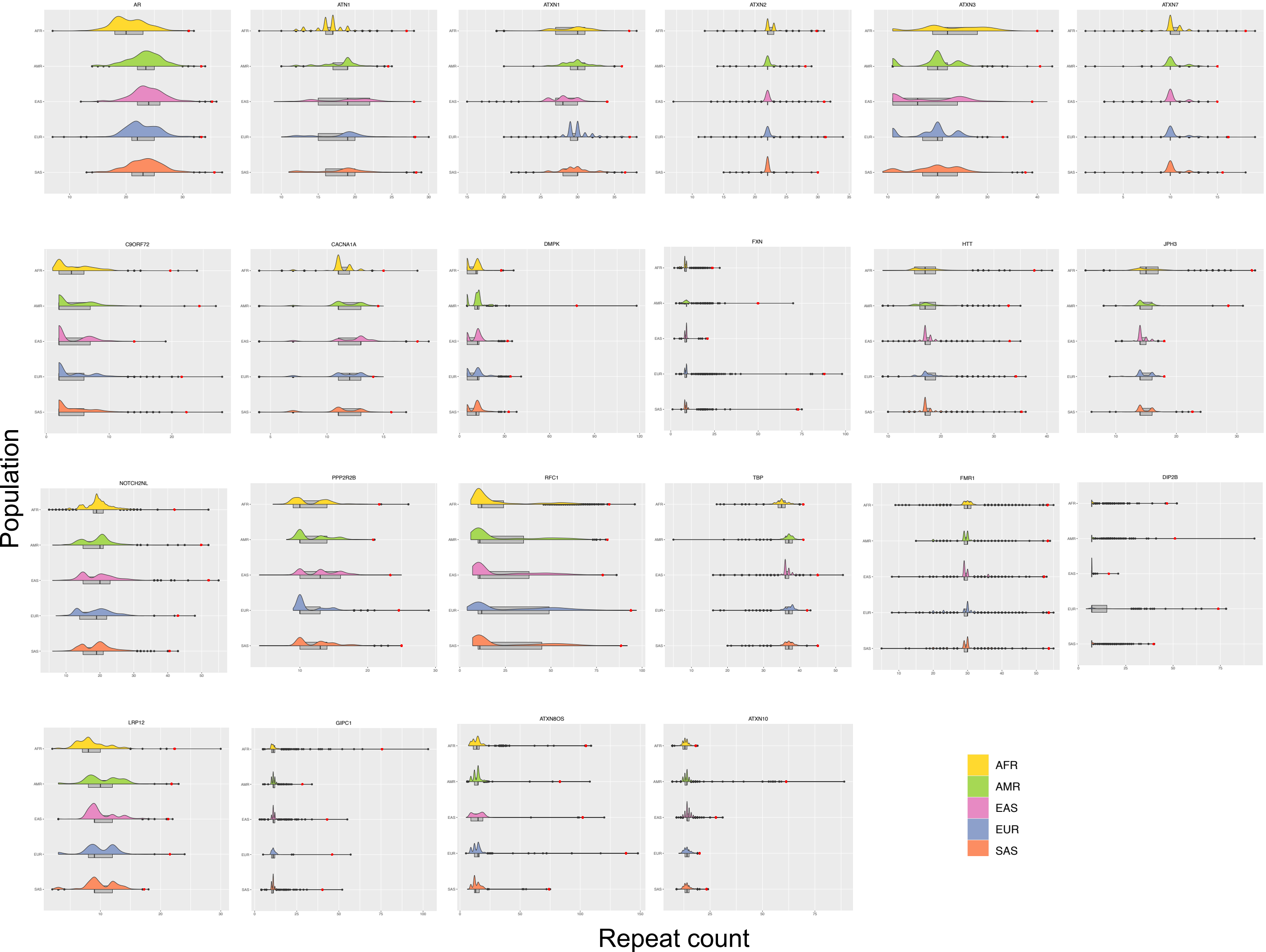
